# Virus detection and identification in minutes using single-particle imaging and deep learning

**DOI:** 10.1101/2020.10.13.20212035

**Authors:** Nicolas Shiaelis, Alexander Tometzki, Leon Peto, Andrew McMahon, Christof Hepp, Erica Bickerton, Cyril Favard, Delphine Muriaux, Monique Andersson, Sarah Oakley, Alison Vaughan, Philippa C. Matthews, Nicole Stoesser, Derrick Crook, Achillefs N. Kapanidis, Nicole C. Robb

## Abstract

The increasing frequency and magnitude of viral outbreaks in recent decades, epitomized by the current COVID-19 pandemic, has resulted in an urgent need for rapid and sensitive diagnostic methods. Here, we present a methodology for virus detection and identification that uses a convolutional neural network to distinguish between microscopy images of single intact particles of different viruses. Our assay achieves labeling, imaging and virus identification in less than five minutes and does not require any lysis, purification or amplification steps. The trained neural network was able to differentiate SARS-CoV-2 from negative clinical samples, as well as from other common respiratory pathogens such as influenza and seasonal human coronaviruses. Additionally, we were able to differentiate closely related strains of influenza, as well as SARS-CoV-2 variants. Single-particle imaging combined with deep learning therefore offers a promising alternative to traditional viral diagnostic and genomic sequencing methods, and has the potential for significant impact.

## INTRODUCTION

The SARS-CoV-2 betacoronavirus has infected hundreds of millions of people since its emergence, resulting in numerous deaths, and causing worldwide social and economic disruption. The emergence of a number of variants of concern (VOCs) that pose an increased risk to global public health by affecting transmission, associated disease severity or vaccine efficacy, have further complicated response efforts.

Current SARS-CoV-2 diagnostic methods include nucleic acid amplification tests, antigen detection, and serology tests (*1*). Reverse transcriptase polymerase chain reaction (RT-PCR) is considered the gold standard for diagnosis; however, RT-PCR takes several hours to provide a result, is restricted to specialized laboratories (as it requires viral lysis and RNA extraction), and can be limited by supply chain issues. Isothermal nucleic acid amplification methods, such as loop-mediated isothermal amplification (RT-LAMP), offer a promising alternative that does not require thermal cycling and can provide results within an hour (*2-7*); however, these methods are still subject to similar supply chain issues as RT-PCR. Lateral-flow immunochromatographic assays using gold nanoparticles and a colorimetric label to detect SARS-CoV-2-specific antigens provide a rapid platform for point-of-contact virus detection, but can have lower sensitivities (*8*). Viral strain, or variant, identification largely relies on sequencing of the viral genome. There is thus an urgent need for new viral detection approaches, particularly ones that can be deployed in non-laboratory settings.

In previous published work we described a robust method to rapidly label enveloped virus particles using a solution of a divalent cation (such as Ca^2+^), short DNAs of non-specific sequence and a particle with a negatively charged lipid bilayer, and suggested that the cations facilitate an interaction between the negatively charged polar heads of the viral lipid membrane and the negatively charged phosphates of the nucleic acid (*9*). By including a fluorophore on the DNAs, we have been able to easily generate bright fluorescent particles for any enveloped virus tested to date (multiple strains of Influenza A, Influenza B, baculovirus, respiratory syncytial virus (RSV), Infectious Bronchitis Virus (IBV), human coronaviruses OC43, HKU1 and NS63, and SARS-CoV-2). We have previously characterised the labelling method to show that virus-specific signals were only observed when the cation, virus and fluorescent DNA were all present (with signals being absent in controls where any one of these major components were excluded), that we could co-stain cation-labelled virus particles with virus-specific antibodies, and that the size of the labelled particles correlated perfectly with that observed in electron microscopy images of the virus (*9*), providing us with confidence that we can specifically label viruses with this method.

To address the need for new viral detection approaches, we have used this labelling method to develop a novel diagnostic test that relies on the detection of intact virus particles using wide-field fluorescence imaging. Our method starts with the near-instantaneous fluorescence labeling of viruses in a sample; we subsequently surface-immobilise labelled particles, collect diffraction-limited images containing thousands of labelled particles, and finally use image analysis and machine-learning to identify different viruses in biological and clinical samples (Fig.1A). Our approach exploits the fact that distinct virus types and strains have differences in surface chemistry, size, and shape, which in turn affects the fluorophore distribution and density over the surface of different viruses. Such differences can be captured by convolutional neural networks (CNNs) (*10, 11*), which have been used previously to classify super-resolved microscopy images of heterogeneous virus populations into particle classes with distinct structural features (*12*), and to detect virus particles in transmission electron microscopy images (*13*). We have shown that we can use this methodology to differentiate a range of viruses in clinical samples, with high overall sample accuracies of 98.0% (using 51 clinical samples on multiple different versions of the trained network) and 97.1% (using 104 clinical samples on a single trained network). We therefore see an opportunity for our testing platform to make an impact not only during pandemics, but also in the future as a general-use method for diagnostics and a surveillance platform for new emerging pathogens.

**Figure 1.**
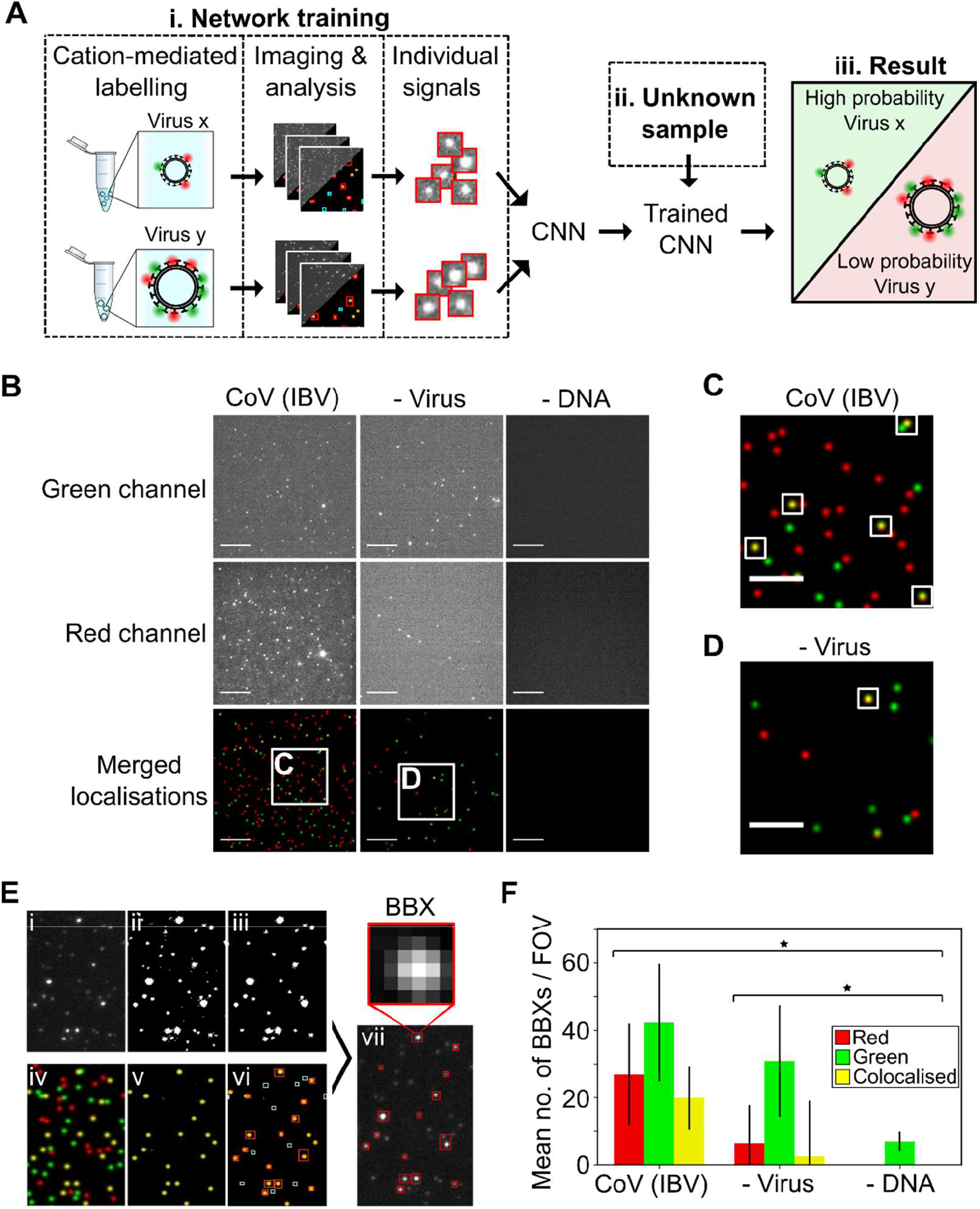
A fluorescent labelling and imaging strategy to detect viruses. A) Overview. i) Viruses were labelled and imaged. Individual signals were isolated and a convolutional neural network (CNN) was trained to exploit differences in the features of different viruses to identify them. ii) Signals from unknown samples can then be fed into the trained CNN to allow iii) virus classification. B) Representative fields of view (FOVs) of infectious bronchitis virus (CoV (IBV)). 1×10^4^ PFU/mL virus was labelled with 0.23 M SrCl_2_, 1 nM Cy3 (green) DNA and 1 nM Atto647N (red) DNA before being imaged. Green DNA was observed in the green channel (top panels) and red DNA in the red channel (middle panels); merged red and green localisations are shown in the lower panels. Scale bar 10µm. A negative control where DNA was replaced with water is included. C&D) Zoomed-in images from B), white boxes represent examples of colocalised particles. Scale bar, 5 µm. E) Segmentation process. (i) Cropped FOV from the red channel. (ii) Intensity filtering applied to i) to produce a binary image. (iii) Area filtering applied to ii) to include only 10-100 pixel objects. (iv) Location image associated with i). (v) Colocalised signals in the location image. (vi) Bounding boxes (BBXs) found from iii) drawn onto v). Non-colocalised objects (cyan) are rejected. (vii) Colocalised objects (red) are drawn over i). Scale bar 10µm. F) Plot of mean number of BBXs per FOV for labelled CoV (IBV) and the negative controls. Error bars represent the standard deviation of 81 FOVs from a single slide. Statistical significance was determined by one-way ANOVA, *P=6.01×10^−22^.

## RESULTS

### Labelled virus particles can be efficiently detected with TIRF microscopy

To demonstrate our ability to label, immobilize, and image virus particles, we initially used infectious bronchitis virus (IBV), an avian coronavirus (CoV). We labelled IBV using a divalent cation (here, Sr^2+^, which performs very similarly to Ca^2+^; see below) and a mixture of green and red fluorescent DNAs (labelled with Cy3 or Atto647N fluorophores, respectively); immobilized particles on a chitosan-coated glass slide; and imaged particles using total-internal-reflection fluorescence microscopy (TIRF) (Sup.Fig.1A). Fluorescent labelling was achieved within seconds via a single-step addition of labelling mixture (see Methods), after which the viruses were immediately immobilized. The resulting images contained particles with either single green or red fluorescence signals (shown as green and red particles), as well as colocalised green and red fluorescence signals (shown as yellow particles) (Fig.1B-D). Efficient virus labelling was achieved using either CaCl_2_ (Sup.Fig.1B&C) or SrCl_2_ (Fig. 1B-D), although both solutions resulted in a number of colocalized signals in the virus-negative controls, likely due to random coincidence or cation-mediated clustering of DNAs on the surface. Omission of DNAs resulted in complete loss of the fluorescent signal (Fig.1B, right panels).

Prior to use for machine learning, the virus images were pre-processed to isolate individual image signals into bounding boxes (BBXs) using segmentation of the (FOV) through adaptive filtering (Fig.1E). The BBX-based approach is preferred over the use of full field of views (FOVs) for classification, since the former is immune to features such as virus concentration or variability in the background and illumination pattern. The raw FOVs from the red channel (Fig.1Ei) were converted into a binary format (Fig.1Eii) and area filtering used to disregard objects with a total area (i.e. width x length) smaller than 10 pixels (single fluorophores) or larger than 100 pixels (aggregates, cells or cell fragments) (Fig.1Eiii). At the same time, to enrich our sampling for viruses and exclude signals not arising from virus particles, the location image (showing the green, red and yellow signals from both channels; Fig.1Eiv) was used to identify colocalised signals (Fig.1Ev). This information was then combined with the signals identified in the filtered binary image (Fig.1Eiii) to reject signals not meeting the colocalisation condition (Fig.1Evi; cyan boxes) and retain signals meeting the colocalisation condition (Fig.1Evi-vii; red boxes). The segmentation was fully automated, allowing each FOV to be processed in ∼2 seconds. In this experiment, the mean number of colocalised BBXs per FOV obtained when IBV was present was ∼6-fold higher than when the virus was absent (Fig.1F), confirming that virus-specific images are being captured in our pre-processing step.

### Positive and negative virus images can be distinguished using deep learning

Having shown that we could efficiently image virus samples and isolate the resulting signals into BBXs, we hypothesized that we could use a custom-built CNN to differentiate between signals observed in virus-positive and virus-negative samples, as well as between images of different viruses. To explore this, we fluorescently labelled and imaged IBV, three laboratory-grown influenza A strains: H3N2 A/Udorn/72 (Udorn), H3N2 A/Aichi/68 (X31), and H1N1 A/PR8/8/34 (PR8), and a virus-negative control consisting of virus-free cell culture media (Fig.2A). The viruses are similar in size and shape, and cannot be distinguished by eye in diffraction-limited microscope images of fluorescently labelled particles (Sup.Fig.2). After image segmentation and examination of the properties of the resulting BBXs however, we observed that the four viruses exhibited small, yet statistically significant, differences in maximum pixel intensity, area, and semi-major-to-semi-minor-axis-ratio within the BBXs (Fig.2B-D); e.g., IBV appears brighter than influenza, whereas Udorn occupies a larger area than the other viruses.

**Figure 2.**
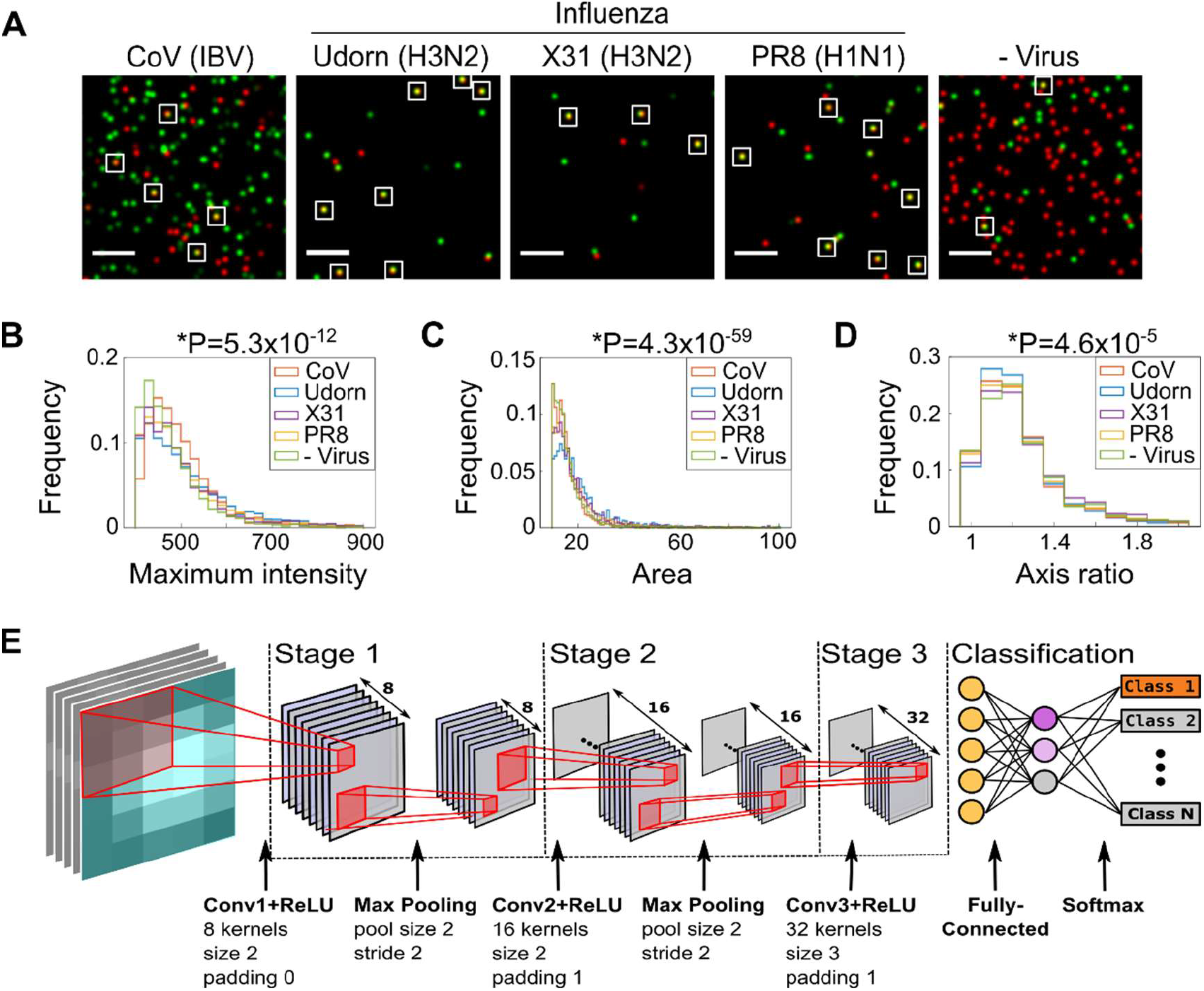
Design of a convolutional neural network to classify imaged viruses. A) Representative FOVs of fluorescently labelled coronavirus (CoV (IBV)), two strains of H3N2 influenza (A/Udorn/72 (Udorn) and A/Aichi/68 (X31)), an H1N1 influenza strain (A/PR8/8/34 (PR8)) and a negative control where virus was substituted with allantoic fluid. The samples were immobilized and labelled with 0.23M SrCl_2_, 1nM Cy3 (green) DNA and 1nM Atto647N (red) DNA before being imaged. Merged red and green localisations are shown, examples of colocalisations are highlighted with white boxes. Scale bar 10µm. B-D) Normalised frequency plots of the maximum pixel intensity, area and semi-major-to-semi-minor-axis-ratio within the BBXs of the four different viruses. Values taken from 81 FOVs from a single slide for each virus. Statistical significance was determined by one-way ANOVA, P values depicted above graphs. E) Illustration of the 15-layer shallow convolutional neural network. Following the input layer (inputs comprising BBXs from the segmentation process), the network consists of three convolutions (stages 1-3). Stages 1 and 2 each contain a ReLU layer to introduce non-linearity, a batch normalisation layer (not shown) and a max pooling layer, while stage 3 lacks a max-pooling layer. The classification stage has a fully-connected layer and a softmax layer to convert the output of the previous layer to a normalised probability distribution, allowing the initial input to be classified.

This was further supported by super-resolution imaging of cation-labelled virus particles. Fluorescence-based super-resolution microscopy allowed us to take both diffraction limited and high resolution images of the same fields of view, providing a direct comparison between the signals isolated into BBXs for the machine learning and their super-resolved versions. We immobilised biotinylated viruses on pegylated coverslips before labelling them with CaCl_2_ and a DNA conjugated to a photoswitchable Alexa647 dye. When imaged, the fluorescent signals from the Alexa647 DNAs on the virus particles were recorded and each resulting localization was precisely fitted to reconstruct a super-resolved image. Cluster analysis of the super-resolved localisations revealed that the fluorescent signals observed in the diffraction limited images of labelled samples correspond to particles of the correct size and shape of virions, and that different virus classes appear to have subtle differences in their labelling density, area and shape (Sup.Fig.3). These small differences, as well as more abstract image features such as pixel correlations, can be exploited by deep learning algorithms to classify the viruses.

To classify the different samples, we constructed a 15-layer CNN (Fig.2E, see legend for detail). We started by imaging IBV and a virus-negative control consisting of only SrCl_2_ and DNA. The two samples were independently imaged four times each over a three-day period. Imaging over three days allowed any potential heterogeneity in the image procurement process (such as small differences in temperature on different days) to be captured, in order to enhance the ability of the trained models to classify data from future datasets. The resulting BBXs obtained for each sample were then randomly divided into a training dataset (70%) and a validation dataset (30%). The training dataset was used to train the CNN to differentiate IBV from negative signals, using ∼3000 BBXs per sample.

The trained network was validated using the remaining 30% of the data (that the network had never seen before). The first data point in the network validation session was at 50% accuracy (as expected for a completely random classification of objects into two categories), followed by an initial rapid increase in validation accuracy as the network detected the most obvious parameters, followed by a slower increase as the number of iterations increased (Sup.Fig.4A). This was accompanied by a similar decrease in the Loss Function (Sup.Fig.4B). The entire training and validation process took 12 minutes to complete (Sup.Fig.4C).

Results of the network validation are shown as a confusion matrix, commonly used to visualize performance measures for classification problems (Fig.3A). The rows correspond to the predicted class (Output Class), the columns to the true class (Target Class), and the far-right, bottom cell represents the overall validation accuracy (hereafter, accuracy) of the model for each classified particle. For IBV vs. negative, the trained network was able to differentiate positive and negative samples with high accuracy (91.4%), sensitivity (91.9%) and specificity (90.9%) (Fig.3B). Of note, these probabilities refer to the identification of single virus particles in the sample and not the whole sample; the probability of correctly identifying a sample with hundreds or thousands of virus particles will therefore increase (see later).

**Figure 3.**
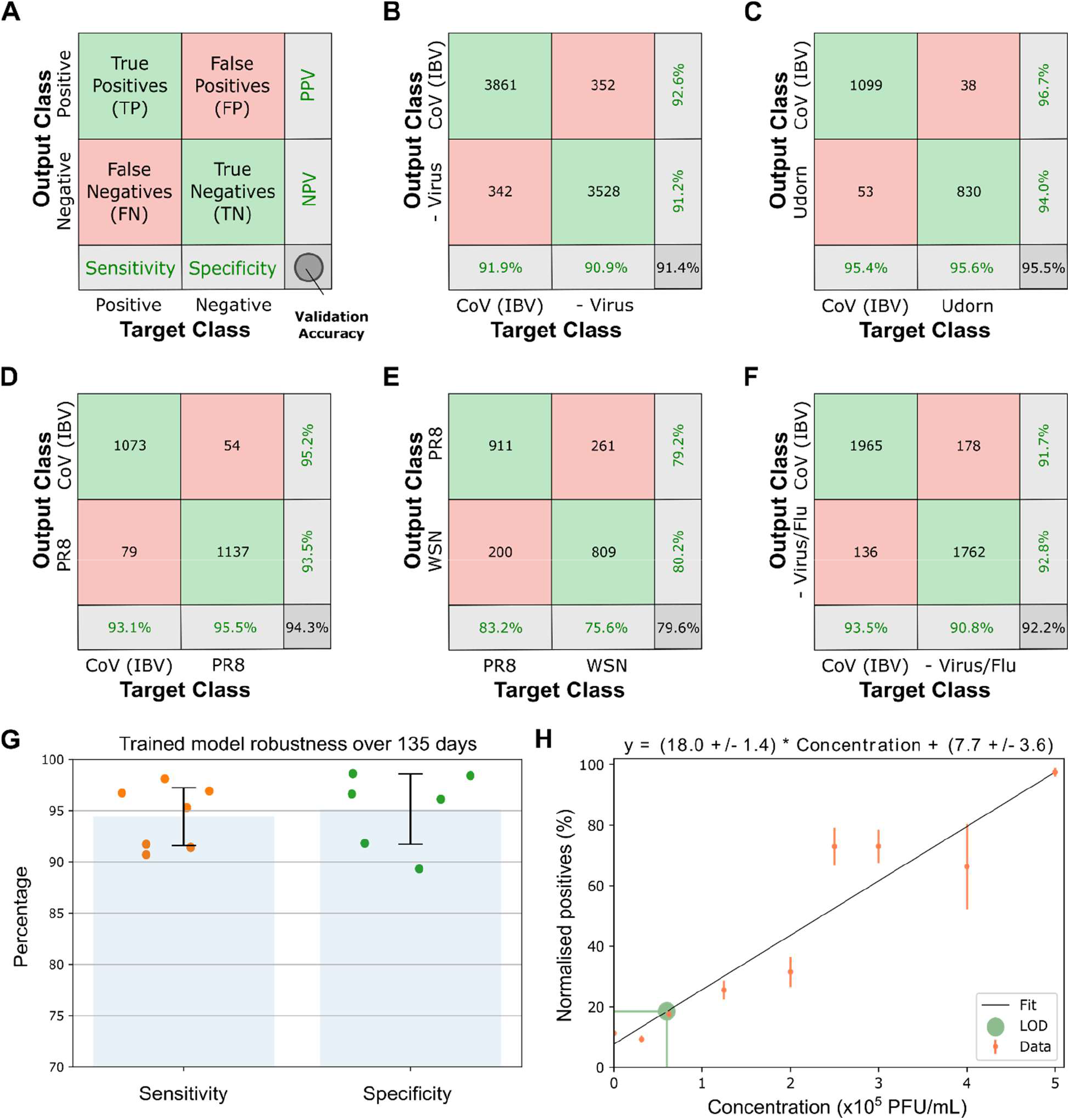
Network validation results for laboratory-grown virus strains. A) Network validation results shown as a confusion matrix. Rows - predicted class (output class); columns - true class (target class); right column - positive and negative predictive values (percentages of BBXs that are correctly and incorrectly predicted); bottom row - sensitivity and specificity. B) Confusion matrix of CoV (IBV) positive and negative samples. C&D) Confusion matrices of CoV (IBV) vs. influenza Udorn or PR8. E) Confusion matrix of influenza PR8 vs. influenza WSN. F) Confusion matrix of CoV (IBV) vs. a pooled dataset of the virus-negative control and three influenza A strains. G) A trained network is robust over significant time. The network was trained on data from images of the virus IBV and allantoic fluid as a negative control. Each data point (orange for sensitivity; green for specificity) corresponds to the classification result for signals detected at different dates over a period of 135 days. Error bars represent standard deviation. H) Defining the limit of detection for accurate machine learning classification. Increasing concentrations of IBV were labelled and imaged, the resulting images were fed into the trained network. The number of normalised positive particles (positive particles/all particles) increased linearly with increasing virus concentration. Error bars represent standard deviation. The limit of detection (LOD) was defined as 6×10^4^ PFU/mL, with 99.85% certainty.

### Efficient classification of different virus strains across optical systems using deep learning

Next, we tested the network’s ability to distinguish between different virus types and strains by training the network on BBXs obtained (as described above) from images of IBV and influenza Udorn, X31, PR8 and H1N1 A/WSN/33 (WSN) strains. The network easily distinguished between the coronavirus and influenza, with a validation accuracy of 95.5% for IBV vs. Udorn (Fig.3C) and 94.3% for IBV vs. PR8 (Fig.3D). The network was also able to differentiate between closely related strains of influenza (WSN vs. PR8), albeit with a slightly lower accuracy of 79.6% (Fig.3E), perhaps reflecting the greater homogeneity between H1N1 strains of the same virus. The ability to distinguish between different influenza viruses that were grown in the same cell line (i.e. WSN and PR8 were both grown in MDCK cells) established that our classification is not host-cell dependent. The network was also able to distinguish between IBV and a pooled dataset consisting of the virus-negative control and three influenza strains (92.2%) (Fig.3F), and importantly, was able to distinguish three viruses from each other in a multi-classifier experiment (the coronavirus IBV and two influenza strains, PR8 and WSN; 81.9%) (Sup.Fig.5).

To demonstrate the general applicability of our approach, we performed similar experiments using a second optical system (a Zeiss Elyra 7 with a 63x objective rather than an ONI Nanoimager with a 100x objective). We compared two strains of influenza (WSN and Udorn) with negative samples lacking virus, and with each other; and found that we were able to distinguish the samples with accuracies ranging from ∼74-78% (Sup.Fig.6), thus establishing that virus classification is independent of the imaging conditions (e.g. exposure, illumination) and microscope type (e.g. magnification, numerical aperture). We also showed that a neural network returned robust results over significant time without requiring re-training, with no decrease in sensitivity or specificity over a period of 135 days (Fig.3G).

We estimated the limit of detection (LOD) of our assay by testing the ability of the network to accurately detect decreasing IBV, WSN and SARS-CoV-2 concentrations (Fig.3H and Sup.Fig.7). Training was performed on laboratory propagated virus samples of known titre, followed by normalisation of the number of BBXs in each class by the total number of BBXs in the sample (to counter variations between samples). Images were analysed by the trained network, and the number of particles classified as positive was fitted with increasing virus concentration, giving estimated LODs of 6×10^4^, 4.6×10^4^ and 5.4×10^4^ PFU/mL for the three virus strains tested. This sensitivity, as expected, was lower than that of amplification-based methods like RT-PCR (∼10^2^ PFU/mL (*14*)), however is still within a clinically useful range; SARS-CoV-2 viral loads have been demonstrated to be between 10^4^ to 10^7^ copies per mL in throat swab and sputum samples (*15*).

### Classification of clinical samples with high accuracy

Having demonstrated our assay on laboratory-grown viruses, we next assessed clinical samples (workflow in Fig.4A). Throat swabs from 33 patients negative for virus (as determined by RT-PCR), or positive for SARS-CoV-2, seasonal hCoVs (OC43, HKU1 or NL63) or human influenza A (as determined by RT-PCR) were inactivated with formaldehyde before being labelled and immobilised (see Methods). Robust and reproducible labelling of viruses in clinical samples was achieved (Sup.Fig.8). Images of the samples captured over three different days were used to train and validate the network to answer a variety of paired questions (e.g. SARS-CoV-2 vs. negative, or SARS-CoV-2 vs. hCoVs; details of clinical samples used for network training and validation are described in Table 1), and similarly to above, the results of the network validation were depicted as confusion matrices.

**Figure 4.**
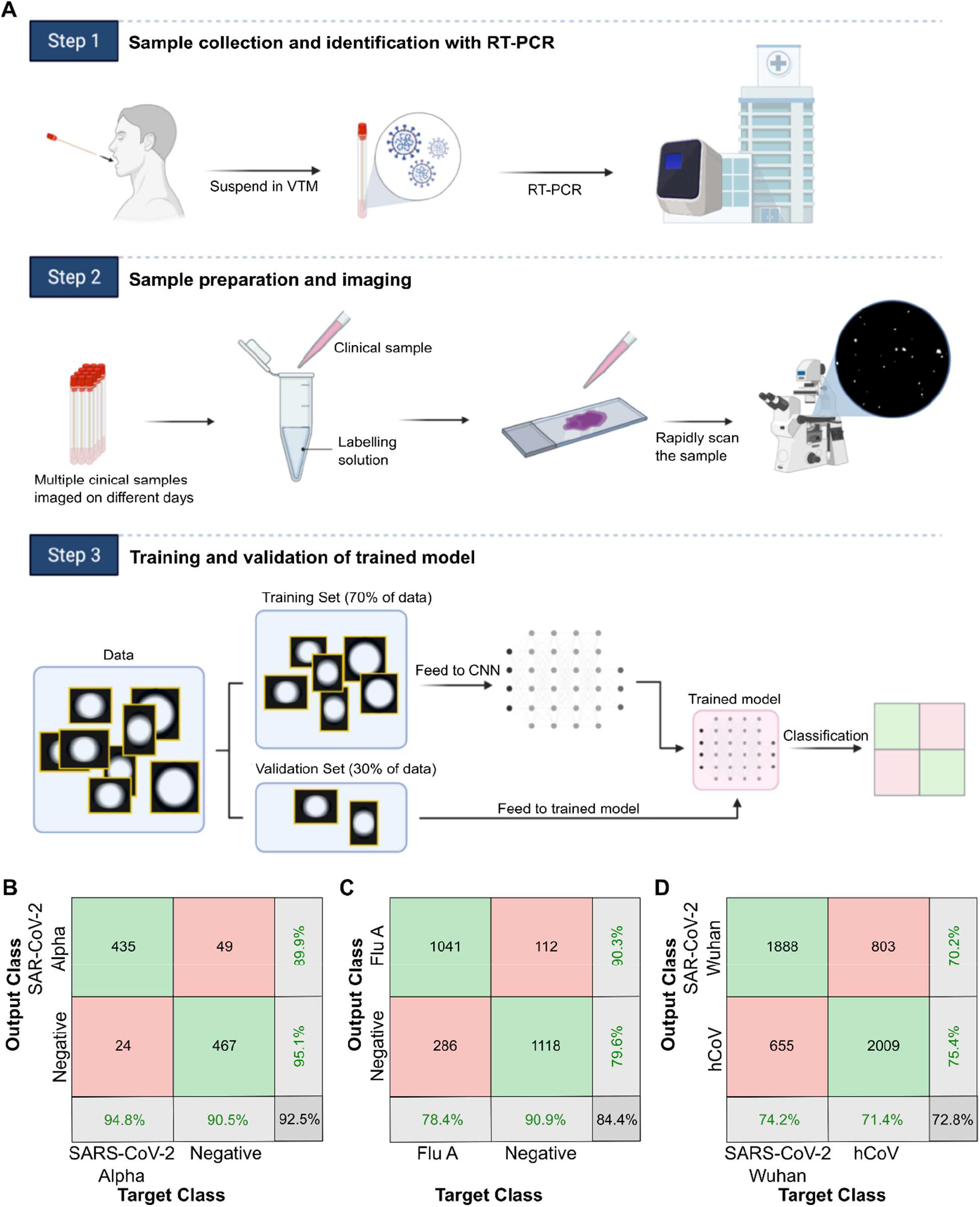
A deep learning network can differentiate viruses in clinical samples. A) Workflow for training and validation of clinical samples. Samples were collected from 33 patients, labelled and imaged on a microscope over three different days. The images were processed to isolate the individual signals into BBXs. 70% of the BBXs were used to train a convolutional neural network (CNN), resulting in a trained model. The remaining 30% of the BBXs were used to validate the trained model, providing the result in a confusion matrix. B) Confusion matrix showing that a trained network could differentiate positive (Alpha variant) SARS-CoV-2 and negative clinical samples. C) Confusion matrix showing that a trained network could differentiate influenza A (Flu A) positive clinical samples from negative samples. D) Confusion matrix showing that a trained network could differentiate SARS-CoV-2 samples (original Wuhan variant) from seasonal human coronavirus (hCoV) samples.

Our initial results with SARS-CoV-2 clinical samples showed a lower validation accuracy than that achieved with laboratory-grown virus strains (∼70% at the BBX level, Sup.Fig.9A). However, the accuracy was substantially improved by performing labelling at a higher pH (pH 8), likely due to the higher isoelectric point (pI; the pH at which the net charge of the particle is zero) of SARS-CoV-2 relative to influenza (pI of ∼9 compared to ∼6) (*16-18*). As the virions are more negatively charged at higher pH, they are more efficiently labelled using the cationic solution and more efficiently captured by the charged chitosan surface on the glass slide, leading to more efficient SARS-CoV-2 detection and improved detection accuracy. Using the optimized protocol, the trained network was able to distinguish between virus-positive and virus-negative clinical samples with excellent accuracy, distinguishing between SARS-CoV-2-positive and negative BBXs with a validation accuracy of ∼93% (Fig.4B).

We could also distinguish between Flu A and negative BBXs with a validation accuracy of ∼84% at the BBX level (Fig.4C), and between seasonal hCoV and negative samples with an accuracy of ∼78% (Sup.Fig.9B). The network could also distinguish SARS-CoV-2 from seasonal hCoVs with a validation accuracy of ∼73% (Fig.4D) and SARS-CoV-2 from Flu A with a validation accuracy of ∼70% (Sup.Fig.9C), potentially useful in diagnosing co-circulating infections. Lastly, the network was able to distinguish between negative samples and combined data from two variants of SARS-CoV-2, the original Wuhan strain (SARS-CoV-2) and the Alpha variant, with an accuracy of ∼75% (Sup.Fig.9D), and between the two variants with an accuracy of∼70% (Sup.Fig.9E).

### Testing of trained networks on independent clinical samples

Next, we tested the trained network’s ability to diagnose independent clinical samples never seen before for either network training or validation. A total of 51 samples (from a different set of patients to those used for network training/validation), comprised of negative samples, or samples positive for SARS-CoV-2, Flu A or seasonal hCoVs, were imaged on a fourth day and assessed by the trained networks described in the section above within a few seconds. By comparing the results to RT-PCR and carrying out chi-squared tests where necessary (Fig.5A and Sup.Fig.10, Steps 1&2), we showed that 50 out of 51 clinical samples tested were classified correctly, giving an excellent overall sample accuracy of 98.0% (Fig.5B and Table 2). For the negative samples, 11 out of 11 were classified correctly, giving a perfect sample specificity of 100%, and for the positive samples, 39 out of 40 were classified correctly, giving a very high sample sensitivity of 97.5%. We observed that the single incorrectly classified sample provided a much lower number of BBXs than the other samples (Table 2), suggesting that the viral load in this sample may have been close our limit of detection and thus explaining the misclassification.

**Figure 5.**
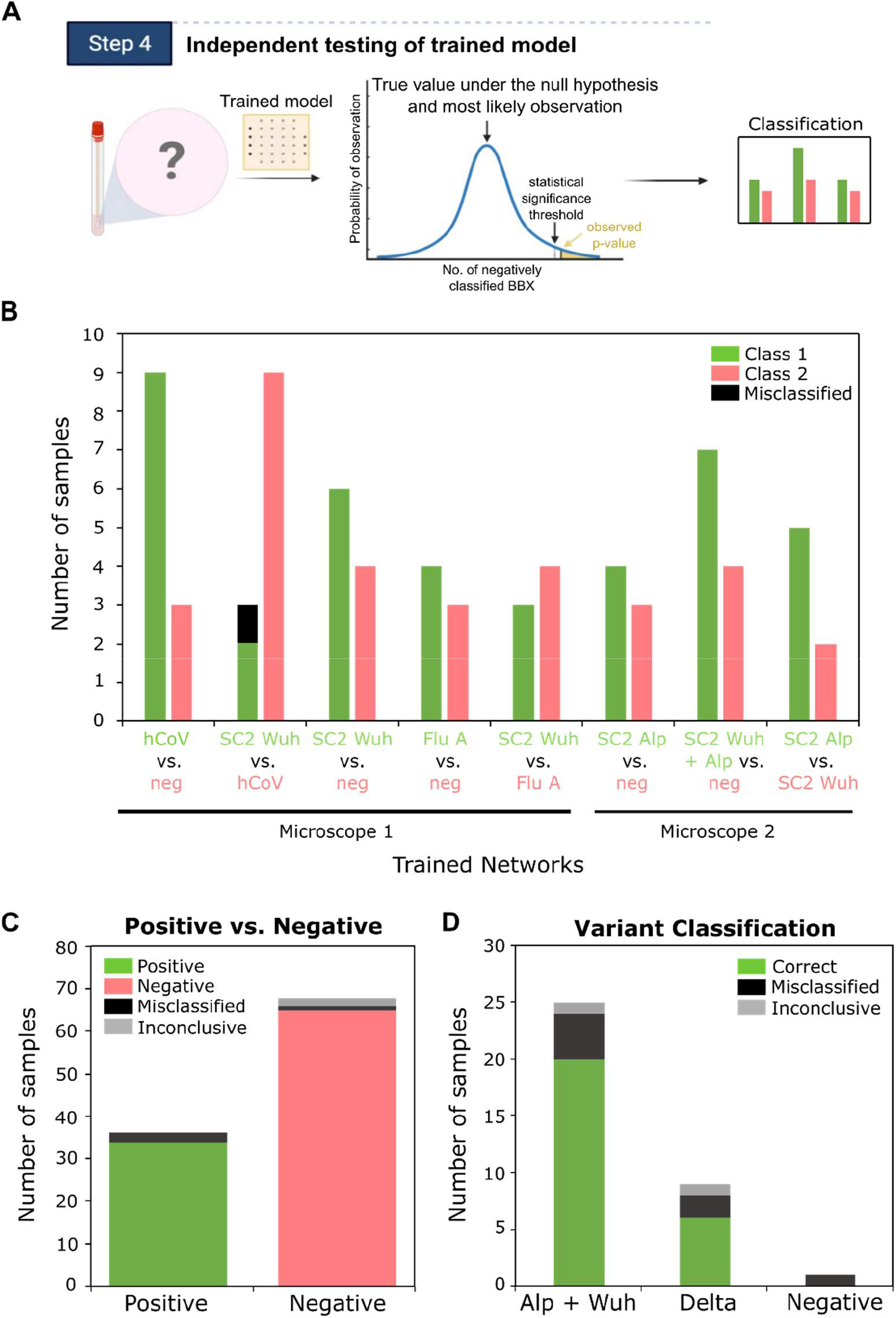
Independent testing of the trained network with clinical samples. A) Schematic of workflow of independent testing. Previously unseen samples are imaged, the images are processed into BBXs which are fed through a trained network. When necessary, a chi-squared statistical test is performed to test the null hypothesis that the sample is negative. If the p-value is smaller than a pre-set confidence threshold the null hypothesis is rejected and the sample is classified as positive. If the p-value is greater than the threshold the sample is classified as negative. B) Summary of independent testing results using multiple trained models. 51 patient samples that were not used for network training or validation were run through different trained versions of the network, detailed on the bottom of the plot. Some samples were tested in multiple versions of the network, for further details see Table 2. Chi-squared tests were carried out to classify the samples (see Sup.Fig.8 and Table 2; samples with a p-value smaller than a pre-set confidence threshold were classified as positive) and the results compared to RT-PCR. 50 out of 51 samples were classified correctly (incorrect classification shown in black), giving an overall sample accuracy of 98.03%. Results were obtained using two different microscopes (see methods). C) Summary of independent testing results using a single trained network. 104 patient samples were analysed as in B), but tested in a single trained network (SARS-CoV-2 vs. negative). D) Variant classification of clinical samples identified as positive in C). The BBXs from images of the clinical samples classified as positive by the first network were passed through a second trained model (Wuhan + Alpha SARS-CoV-2 vs. Delta SARS-CoV-2).

Within this analysis, we also tested whether we could differentiate different SARS-CoV-2 variants. The BBXs from images of seven clinical samples that had been classified as positive by a SARS-CoV-2 vs. negative network were passed through a second trained network to test whether it was the original Wuhan variant or the Alpha variant that first arose in the UK in 2020 (Sup.Fig.10, Steps 3&4). All seven variant samples were classified correctly (Fig.5B).

Given the potential clinical relevance of a test that can diagnose different SARS-CoV-2 variants without the need for sequencing, we decided to explore this further using an additional 104 clinical samples, 68 of which were determined to be virus-negative and 36 of which were determined to be SARS-CoV-2 positive by RT-PCR. The samples were taken from patients from November 2020 to July 2021, and of the positive samples, 14 were the original Wuhan variant, 12 were the Alpha variant (as suggested by a spike gene target failure in RT-PCR) and 10 were the Delta variant. Samples were labelled, immobilised and imaged as normal, followed by processing of the images into BBXs. In the first step of the analysis the 104 samples were classified as either SARS-CoV-2-positive or negative (results of the network validation in Sup.Fig.11A). Two of the negative samples were inconclusive due to a low number of BBXs (less than the 5 needed for the chi-squared test). Of the remaining 102 samples, all but three of the samples were classified correctly (Fig.5C). Of the three samples that were misclassified one was a negative sample and 2 were positive samples (Fig.5C), providing us with an overall sample accuracy of 97.1%, a sensitivity of 94.4% and a specificity of 98.5%.

For the subsequent variant classification step, the BBXs from images of the clinical samples classified as positive by the first network (35 samples in total) were passed through a second trained model (Wuhan + Alpha SARS-CoV-2 vs. Delta SARS-CoV-2) (Sup.Fig.11B). Two samples gave inconclusive results as the resulting p-values were closer than 3 orders of magnitude (Sup.Fig.10, Steps 3&4). The single negative sample that had been incorrectly classified as positive by the first trained model was classified as Delta, while six remaining positive samples were misclassified (two Delta, two Alpha and 2 Wuhan variant), giving an overall sample accuracy of 77.1%, inconclusive rate of 5.7% and misclassification rate of 17.1% (Fig.5D and Table 4). Whilst our variant classification accuracy is lower than our overall positive vs. negative test accuracy, we believe that this may still be of use in the context of a rapid variant screening assay in the absence of sequencing facilities.

## DISCUSSION

In summary, we have shown a proof of principle for the use of single-particle fluorescence microscopy and deep learning to rapidly detect and classify viruses, including coronaviruses. We have carried out two clinical tests of our method using 155 patient samples in total, which provided high overall sample accuracies of 98.0% and 97.1%. We note that these results were obtained with samples collected over a significant time-period, using a range of different collection kits containing different volumes of viral transport media, and stored at different temperatures for varying periods of time. Given these sampling inconsistencies, which could potentially impact the number of intact virus particles in each sample, the results of these small-scale clinical trials are extremely encouraging, demonstrating the potential of our method as a viable diagnostic test.

Our initial proof of principle experiments, carried out using viruses grown in cell culture, demonstrated that the CNN can distinguish not only between samples with and without virus, but also between the avian coronavirus IBV and various strains of influenza with high accuracies of >90% per particle (e.g. IBV vs. Udorn, IBV vs. PR8). We confirmed our approach using multiple stocks of cell grown viruses, and different microscopes. A truly independent validation of the trained network using samples that have not been used in either training or validation wasn’t possible using cell grown viruses however, as all stocks are essentially the same virus, grown in the same cell line and similar conditions, even when grown at different times. Importantly, this was possible using clinical samples though, as novel samples could be obtained from new patients that were entirely independent of those used for training/validation. The accurate classification of completely independent clinical samples served as clear proof that our network could use image information to accurately classify samples never seen before.

We also accounted for any other potential confounding factors such as sample preparation (by only comparing samples prepared and inactivated in the same way), as well as differences in virus concentration and differences in imaging quality, by only classifying the isolated signals from individual viruses and not full fields-of-view. This approach renders the network completely agnostic to virus concentration, signal density or small day-to-day imaging differences such as uneven illumination, and means that virus classification is solely dependent on the fluorescent images of the virus. We have even shown that related H3N2 virus strains, prepared in the same way in the same cells, could be reliably distinguished from each other, establishing that classification is independent of the host cell that the virus samples were grown in. All of this, together with our finding that clinical samples that produced different CT values in RT-PCR were correctly classified independently of their concentration, provides several lines of strong evidence that our network can truly differentiate between different viruses.

The power of our method comes from the ability to rapidly and universally label enveloped viruses in a sample, and swiftly image them using diffraction limited microscopy. We have shown that even with the limited information present in the low-resolution images, a trained CNN can very effectively differentiate between virus strains. This is based on our findings that different virus families, and even different virus strains, exhibit small differences in their distributions of size, shape and labelling efficiency when labelled using the cation mediated method (Fig.2 and Sup.Fig.3). The labelling is an electrostatic interaction between the phosphate backbone of the DNA and the lipid membrane of the virus (*9*). Different viruses will therefore exhibit differences in labelling efficiency and coverage due to their different isoelectric points (the pH at which a virus has a neutral surface charge), e.g. WSN: 4.7, PR8: 5.3, IBV: 7.2 and SC2: 8.5 (*16-19*). The surface charge of virus strains is also likely to be affected by mutations in the surface glycoproteins, thus explaining our ability to effectively differentiate variants. We explored the patterns ‘seen’ by two different networks using the DeepDreamImage function in Matlab (*20*), which suggests that a network trained to differentiate between negative and positive SARS-CoV-2 samples may see positive signals as rounded shapes with high intensity in the centre of the BBX, whereas the pixels have much lower values in the centre of the BBX for a negative sample, supporting our theory that virus signals are brighter than the signals observed in a negative sample (Sup.Fig.12A&B). Comparison of the patterns observed by a network trained to differentiate between two variants of SARS-CoV-2 are harder to interpret, however there are higher intensity pixels in both than in the negative sample (Sup.Fig.12C&D).

During our second clinical validation that initially tested for SARS-CoV-2-positive or negative, followed by variant classification, two samples did not provide enough BBXs for us to accurately conclude a result using the chi-squared test. Interestingly, both of the inconclusive samples were negatives (validated by RT-PCR); further testing of more samples may establish that this can happen only for negative samples in which case it may be a useful indicator for a negative diagnosis. The small number of positive samples that were misclassified may have been due to low viral load, which can be improved through further optimisation of the protocol, such as by improving virus immobilisation, sample concentration, or limiting the time the sample is stored for before being imaged. It is possible that the single negative sample that was misclassified may have contained another virus that was not SARS-CoV-2; in further iterations of the test we will use multi-classifier networks to recognise all the major families of circulating respiratory viruses (e.g., Influenza, HCoVs, SARS-CoV-2 and RSV), providing a multi-pathogen testing platform.

The current gold standard for viral diagnostics is RT-PCR, which requires a time consuming (∼30 mins) RNA extraction step, followed by the main assay (which can take several hours). In this manuscript, we describe a laboratory-based proof-of-principle assay that involves i) instantaneous sample labelling, ii) 10 sec for sample mounting, iii) 40 sec for focusing, iv) 2 min for image acquisition (81 FOVs) and v) 20 sec for analysis, thus easily providing a result within just 5 minutes. Our assay requires no RNA extraction but in order to easily work with clinical samples in a containment level 2 laboratory, we initially inactivated samples with a low concentration of formaldehyde (4%) for 30 minutes prior to sample preparation, which has been shown to maintain virus particle shape while rendering them non-infectious (*21*). In later experiments presented here, we moved to working with samples inactivated with 1% formaldehyde in just 5 minutes (having used plaque assays to show that this lower concentration and time was still sufficient to fully inactivate samples); rendering the entire test complete from start to finish within just 10 minutes. This makes our test significantly faster than RT-PCR, which may render the test invaluable in point-of-care situations where time is critical. Our envisaged commercial version of the test will not require inactivation at all (making it even faster) and will use a cost-effective and simplified microscope, making the assay accessible to many.

Our approach also avoids the need for viral lysis or amplification and the associated cost, tedium and supply-chain issues. A key advantage of our test over enzyme-based methods such as RT-PCR or RT-LAMP is that the reagents are very affordable. The most expensive component is the fluorescent DNA, which we typically use at 1 nM concentration in a very small sample volume (20 μL); even in the small volumes that we order DNA, the cost amounts to ∼2.7 pence to run 10,000 labelling reactions, demonstrating the scalability of our test. The non-specific detection of intact viral particles (rather than genome fragments) can report directly on infectivity (although this was not evaluated here), and has the advantages of speed, the ability to detect multiple virus types in a single labelling step, and robustness against potential mutations in the viral genome. Our algorithms are versatile and can be trained to differentiate between many different viruses, independently of how they are labelled, immobilized and imaged.

Here, we have tested the ability of our models to differentiate between the original genotype of SARS-CoV-2, as well as two variants of concern, Alpha and Delta. The ease with which the network can be retrained suggests that it can be rapidly repurposed to detect new and emerging SARS-CoV-2 variants such as Omicron. Given its simplicity and rapid nature, our technology could also be used outside of specialized laboratories, such as in airports, workplaces and care homes. These unique capabilities should enable extremely rapid, mobile, and real-time analysis of patient and community samples during pandemic situations.

## METHODS

### Laboratory grown virus strains and DNAs

The influenza strains (H1N1 A/Puerto Rico/8/1934 (PR8), H3N2 A/Udorn/72 (Udorn), H1N1 A/WSN/33 (WSN) and H3N2 A/Aichi/68 (X31)) used in this study have been described previously (*9*). Briefly, WSN, PR8 and Udorn were grown in Madin-Darby bovine kidney (MDBK) or Madin-Darby canine kidney (MDCK) cells and X31 was grown in embryonated chicken eggs. The cell culture supernatant or allantoic fluid was collected and the viruses were titred by plaque assay. Titres of PR8, Udorn, WSN and X31 were 1×10^8^ plaque forming units (PFU)/mL, 1×10^7^ PFU/ mL, 2×10^6^ PFU/mL and 4.5×10^8^ PFU/mL respectively. The coronavirus IBV (Beau-R strain) (*22*) was grown in embryonated chicken eggs and titred by plaque assay (1×10^6^ PFU/mL). Influenza and IBV were inactivated by the addition of 2% formaldehyde before use. SARS-CoV-2 was grown in Vero E6 cells and titred by plaque assay (1.05×10^6^ PFU/mL). The virus was inactivated by addition of 4% formaldehyde before use.

Single-stranded oligonucleotides labelled with either red or green dyes were purchased from IBA (Germany). We have previously characterised a range of DNAs of different length and sequence and shown that as long as the DNA is longer than 20 bases robust labelling occurs, regardless of sequence(*9*). The ‘red’ DNA used in this manuscript was modified at the 5’ end with ATTO647N (5’ ACAGCACCACAGACCACCCGCGGATGCCGGTCCCTACGCGTCGCTGTCACGCTGGCTGTTTGTCTTCCTGCC 3’) and the ‘green’ DNA was modified at the 3’ end with Cy3 (5’ GGGTTTGGGTTGGGTTGGGTTTTTGGGTTTGGGTTGGGTTGGGAAAAA 3’). The DNA used for super-resolution imaging was modified at the 5’ end with Alexa647 (5’ TCCGCTCTCACAATTCCACACATTATACGAGCCGAAGCATAAAGTGTCAAGCCT 3’).

### Clinical samples

Ethical approval was obtained for the use of anonymised oro- or nasopharyngeal specimens from patients for the diagnosis of influenza and other respiratory pathogens, including SARS-CoV-2 (North West-Greater Manchester South Research Ethics Committee [REC], REC Ref:19/NW/0730). Specimens were maintained in Copan Universal Transport Medium (UTM) before being inactivated in 4% final concentration of formaldehyde (Pierce) for 30 minutes at room temperature, or 1% formaldehyde for 5 minutes at room temperature for the 104 samples used in the second clinical trial (*21*). Samples were confirmed as SARS-CoV-2-positive or negative using either the Public Health England 2019-nCoV real-time RT-PCR RdRp gene assay or RealStar SARS-CoV-2 RT-PCR Kit (Altona diagnostics). Testing for other respiratory pathogens and sub-typing of seasonal human coronavirus (hCoV) samples as OC43, HKU1 or NL63 strains was conducted using the BioFire FilmArray Respiratory Panel (Biomerieux, Marcy-L’Etoile, France) and Cepheid Xpert Xpress Flu/RSV (Cepheid, Sunnyvale, CA, USA).

We used 213 clinical samples in total, taken from patients from November 2020 to July 2021. In order to train the network, we imaged samples from different patients over three days (different sample prep from the same patient samples on each day). We used 70% of the BBXs isolated from all the images taken over the three days to train the network, leaving the remaining 30% of the BBXs for network validation, the results of which are shown in the confusion matrices. Each confusion matrix corresponds to an individually trained model. In total, 58 clinical samples were used for training and validation of the network.

We then carried out two independent tests of the trained networks using clinical samples not used for either training or validation. The first test used 51 patient samples comprised of negative samples, or samples positive for SARS-CoV-2, Flu A or seasonal hCoVs. The second test used 104 patient samples comprised of negative samples, or samples positive for SARS-CoV-2. Of the positives, 14 were the original Wuhan variant, 12 were the Alpha variant (indicated by a spike gene target failure in RT-PCR [TaqPath Covi-19 combo kit, ThermoFisher]) (*23*) and 10 were the Delta variant (confirmed through RT-PCR [Taqman SARS-CoV-2 mutation panel [ThermoFisher]).

### Sample preparation

Both positive and negative samples prepared in the same way (e.g. inactivated in the same concentration of formaldehyde or labelled in the same buffer), and only samples similarly prepared were compared with each other. Glass slides were treated with 0.015 mg/mL chitosan (a linear polysaccharide) in 0.1 M acetic acid for 30 min before being washed thrice with MilliQ water, or with 0.01% poly-L-lysine (Sigma) for 15 min (Fig.3A, Sup.Fig.6, Sup.Fig.10C). Unless otherwise stated, virus stocks (typically 10 µL) were diluted in 0.23 M CaCl_2_ or SrCl_2_ (as described in the figure legends) and 1 nM of each fluorescently-labelled DNA in a final volume of 20 µL, before being added to the slide surface. For SARS-CoV-2 imaging, the cationic labelling solution was buffered with 20 mM Tris, pH 8. Virus labelling with CaCl_2_ has been described previously (*9*); SrCl_2_ provides similar results (Fig.1). For laboratory grown virus stocks, negatives were taken using virus-free Minimal Essential Media (Gibco) or allantoic fluid from uninfected eggs in place of the virus.

### Imaging

Images were captured using three wide-field Nanoimager microscopes (*9*). ‘Microscope 1’ was equipped with a Hamamatsu Flash 4 LT.1 sCMOS camera and ‘Microscopes 2 and 3’ were equipped with a Hamamatsu Flash4 V3 sCMOS camera; in all other respects, the systems were identical. The sample was imaged using total internal reflection fluorescence (TIRF) microscopy and a 100x oil-immersion objective. The laser illumination was focused at a typical angle of 53° with respect to the normal. Movies of 5 frames per field of view (FOV) (measuring 75 × 49 µm) were taken at a frequency of 33 Hz and exposure time of 30 ms, with laser intensities kept constant at 0.78 kW/cm^2^ for the red (640 nm) and 1.09 kW/cm^2^ for the green (532 nm) laser. To automate the task and ensure no bias in the selection of FOVs, the whole sample was scanned using the multiple acquisition capability of the microscope; 81 FOVs were imaged in 2 minutes. Defocusing was carefully controlled using an automated autofocus mechanism to bring the sample to a pre-defined axial position before each field of view was exposed to the excitation lasers. This was achieved by imaging the reflection of a near-IR laser off the glass/sample medium interface and matching the image to a pre-recorded reference image.

Data in Sup. Fig. 6 was acquired using a Zeiss Elyra 7 microscope equipped with two pco.edge sCMOS (version 4.2 CL HS) cameras. TIRF images were acquired using the alpha Plan-Apochromat 63x/1.46 oil objective. A laser intensity of 10% for the 641nm laser was used for imaging Atto647N. Laser intensities of 6% for the 561nm and 3% for the 488nm laser were used for imaging Cy3. The exposure time was 50ms.

### Super resolution

For super resolution imaging, passivated microscope slides were prepared by washing in acetone and Vectabond solution (Vector Laboratories) before being incubated with NHS-PEG:Biotin-NHS-PEG in an 80:1 ratio. 0.5 mg/mL neutravidin was incubated for 10 minutes at room temperature on the slide shortly before virus was added. Viruses were biotinylated by incubation in a 1 mg/mL Sulfo-NHS-LC-Biotin (ThermoFisher) for 3 hours at 37°C before being labelled with 0.23 M CaCl_2_ and 1 nM Alexa647-labelled DNA in a final volume of 20 µL, before being added to the slide surface. After incubation for 30 minutes at room temperature the slide was washed thrice in 1x PBS before imaging in 50 mM MEA and an enzymatic oxygen scavenging system consisting of 1 mg/mL glucose oxidase, 40 μg/mL catalase, and 1.0% (wt/vol) glucose. Super-resolution localisations were extracted using the inbuilt Nanoimager software and analysed further in Matlab. Localisations were clustered with DBScan using a minimum cluster size of 50 and an epsilon of 30nm, followed by computing the convex hull to find the area of the clustered points.

### Data Segmentation

Each FOV in the red channel was turned into a binary image using MATLAB’s built-in imbinarize function with adaptive filtering sensitivity set to 0.5. Adaptive filtering uses statistics about the neighbourhood of each pixel it operates on to determine whether the pixel is foreground or background. The filter sensitivity is a variable which, when increased, makes it easier to pass the foreground threshold. The bwpropfilt function was used to exclude objects with an area outside the range 10-100 pixels (1 pixel = 117nm), aiming to disregard free ssDNA and aggregates. We imaged single fluorophores and found that that they did not exit 10 pixels in area, giving us a lower limit, and arbitrarily chose 100 pixels as the upper limit to exclude very large aggregates or cellular debris. The regionprops function was employed to extract properties of each found object: area, semi-major to semi-minor axis ratio (or simply, axis ratio), coordinates of the object’s centre, bounding box (BBX) encasing the object, and maximum pixel intensity within the BBX.

Accompanying each FOV is a location image (LI) summarising the locations of signals received from each channel (red and green); colocalised signals in the LI image were shown in yellow. Objects found in the red FOV were compared with their corresponding signal in the associated LI. Objects that did not arise from colocalised signals were rejected. The qualifying BBXs were then drawn onto the raw FOV and images of the encased individual viruses were saved.

### Machine Learning

The CNN used only the red channel as input, as analysis using both channels was not found to improve the overall accuracy. No normalisation of the images was carried out, however the bounding boxes (BBXs) from the data segmentation had variable sizes (never larger than 17 pixels in any direction due to the size filtering). Thus, all the BBX were resized such that they had a final size of 17×17 pixels by means of padding (adding extra pixels with 0 grey-value until they reach the required size).

The resized images were used as the input for the 15-layer CNN. The network was built using Matlab 2020b and trained using the computer’s GPU (specifications: NVIDIA 2080Ti, 32 GB RAM, i7 processor). The network had 3 convolutional layers in total, with kernels of 2×2 for the first two convolutions and 3×3 for the last one. The learning rate was set to 0.01 and the learning schedule rate remained constant throughout the training. The hyperparameters remained the same throughout the training process for all models; the mini batch size was set to 1000, the maximum number of epochs to 100 and the validation frequency to 20 iterations.

In the classification layer, trainNetwork took the values from the softmax function and assigned each input to one of K mutually exclusive classes using the cross entropy function for a 1-of-K coding scheme (*24*),

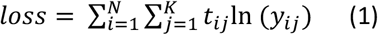

where N is the number of samples, K is the number of classes, *t*_*ij*_ is the indicator that the *i*^*th*^ sample belongs to the *j*^*th*^ class, and *y*_*ij*_ is the output for sample i for class j, which in this case, is the value from the softmax function. That is, it is the probability that the network associates the *i*^*th*^ input with class j (*25*). A stochastic gradient descent with momentum set to 0.9 was used as the optimizer.

### Statistical Analysis

#### Confusion matrices

The results of each network validation are shown as a confusion matrix, which make used of the following terms:

- True positive (TP): BBXs correctly identified as positive,
- False Positive (FP): BBXs incorrectly identified as positive,
- True negative (TN): BBXs correctly identified as negative, and
- False negative (FN): BBXs incorrectly identified as negative.

Sensitivity refers to the ability of the test to correctly identify positive BBXs. It can be calculated by dividing the number of true positives over the total number of positives(*26*).

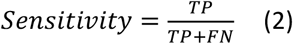

Specificity refers to the ability of the test to correctly identify negative BBXs. It can be calculated by dividing the number of true negatives over the total number of negatives(*26*).

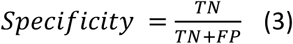

The percentages of BBXs that are correctly and incorrectly predicted by the trained model are known as the positive predictive value (PPV) and negative predictive value (NPV) respectively.

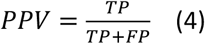

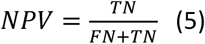

The overall balanced validation accuracy of the model is given by:

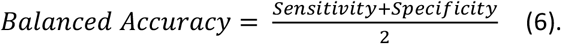

#### Limit of Detection

In order to calculate the limit of detection (LOD), increasing concentrations of the CoV IBV (dilutions in allantoic fluid) were labelled and imaged, the resulting images were pre-processed, and the individual signals were fed into the trained network. The normalised average of *TP (TP/TP+FP*) and standard deviation (STD) were calculated and plotted against the corresponding concentrations as a scatter plot. The plot was fitted as a linear regression, as given by:

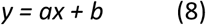

Where the virus concentration was treated as the independent variable and *a* represents the LOD. For the final value of the LOD *a+(3STD)=6*10^4 PFU/mL* was used, which corresponds to a 99.85% confidence interval assuming a normal distribution. Experiments with the influenza strain AWSN/33 were carried out in a similar way.

In order to calculate the LOD of SARS-CoV-2, experiments were carried out in a similar way, however the plot was fitted as a sigmoid, as given by:

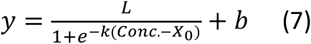

Where L is the maximum normalised positive value (considered as the sensitivity of the model), b adds bias to the output and changes its range from [0,L] to [b,L+b], k scales the input, and X_0_ is the point at which the sigmoid should output the value L/2.

#### Chi-Squared Test

In order to go from single BBX classification to calling the result of a clinical sample as a whole the Chi-squared test was used, which takes into consideration the total number of bounding boxes, the number of BBXs that were classified as positive or negative, and the specificity of the trained model (i.e. the probability of classifying a negative signal as such). By taking into account the specificity of each trained model and the total number of signals in a sample we account for the variability in the number of detected signals between samples. The test also considers that statistically a number of the bounding boxes will be misclassified. The Chi-squared test is a statistical hypothesis test that assumes (the null hypothesis) that the observed frequencies for a categorical variable match the expected frequencies for the categorical variable and can be calculated from the equation below:

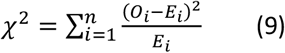

where *χ*^2^= chi squared, *O*_*i*_=observed value, *E*_*i*_=expected value and *n* is the number of labels. The threshold p-value for a test can vary depending on the trained model but in general is smaller than p-value=0.01 which corresponds to a confidence of greater than 99%. In this manuscript the Null hypothesis is that the sample is negative and it is only rejected when the p-value is below the threshold in which case the sample is classified as positive.

## Data Availability

Data available upon request.

## Acknowledgements

We are grateful to Micron Oxford, funded by Wellcome Strategic Awards (091911 and 107457; PI Ilan Davis), for their loan of their microscope and to Nadia Halidi for her help with the instrument. Special thanks to Dr Rebecca Moore and Prof William James (Sir William Dunn School of Pathology, Oxford, UK) for providing laboratory grown SARS-CoV-2. We thank the microbiology laboratory staff at the John Radcliffe Hospital, Oxford. This research was supported by a Royal Society Dorothy Hodgkin Research Fellowship DKR00620 and Research Grant for Research Fellows RGF\R1\180054 (N.C.R.), the University of Oxford COVID-19 Research Response Fund (N.C.R and A.N.K.), a BBSRC-funded studentship (N.S.), and Wellcome Trust grant 110164/Z/15/Z (A.N.K.). All data needed to evaluate the conclusions in the paper are present in the paper and/or the Supplementary Materials.

## Author Contributions

N.S., A.T., A.M., C.H., A.N.K. and N.C.R designed and carried out experiments, analysed data and interpreted results. N.S. wrote analysis software. E.B. provided reagents. C.F. and D.M. validated the virus inactivation method. L.P., M.A., S.O., A.V., P.C.M., N.S. and D.C. collected, diagnosed and typed clinical samples that were considered to be appropriate for the purposes of this study. N.C.R. wrote the manuscript. All authors reviewed and approved the final manuscript.

