## Supplemental data for "Virus detection and identification in minutes using single-particle imaging and deep learning"

**Supplementary Material**

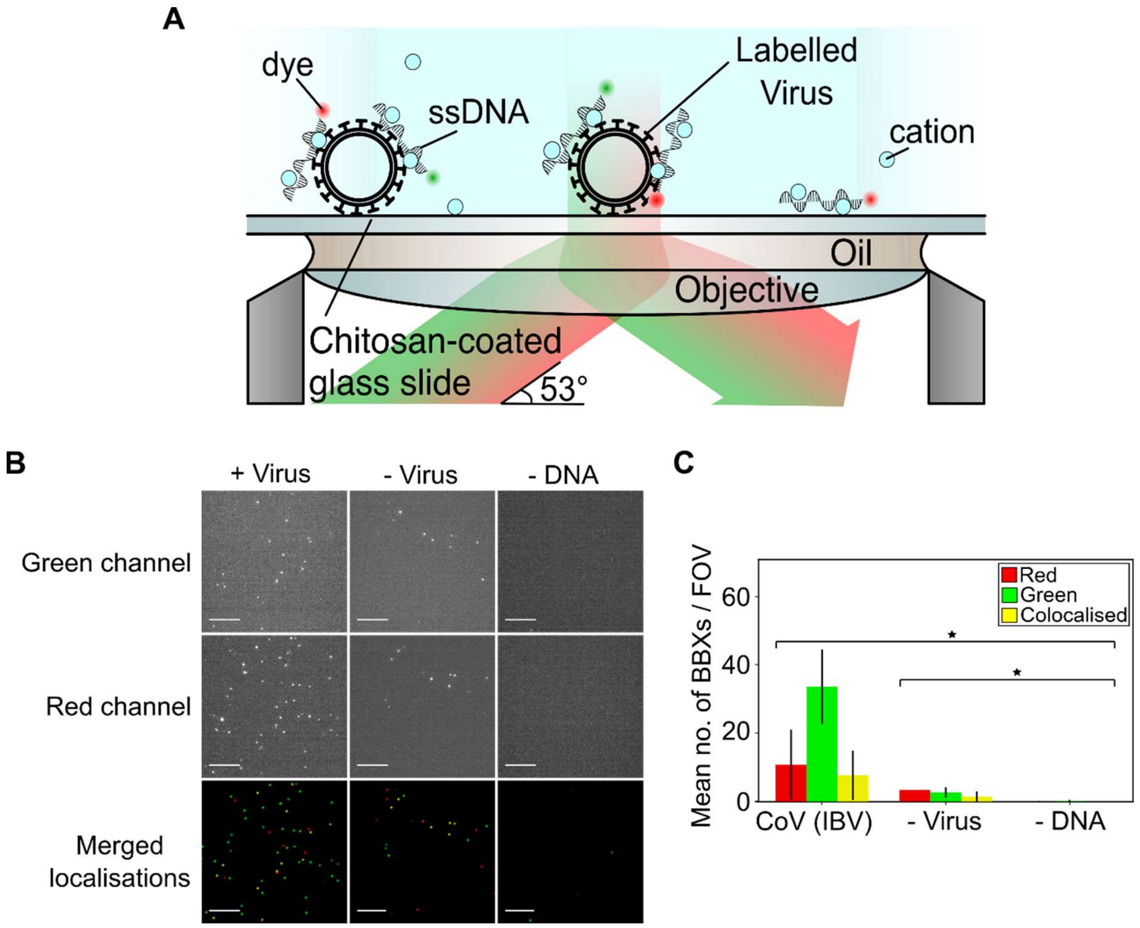


**Sup. Fig. 1. Schematic of virus labelling and immobilisation strategy.** Positively charged cations (e.g. calcium, strontium) bridge the lipid membrane of the virus and the negatively charged phosphate groups on the ssDNA, binding fluorescently labelled ssDNA to the surface of the virus. Labelled viruses were immobilised on a chitosan-coated glass slide and illuminated with red and green laser light on a widefield Total Internal Reflection Fluorescence (TIRF) microscope. B) Representative fields of view (FOVs) of fluorescently labelled infectious bronchitis virus (CoV (IBV)). The virus sample, at a final concentration of $1x10^4$ PFU/mL, was immobilized and labelled with 0.23 M $CaCl_2$, 1 nM Cy3 (green) DNA and 1 nM Atto647N (red) DNA before being imaged. Green DNA was observed in the green channel (top panels) and red DNA in the red channel (middle panels); merged red and green localisations are shown in the lower panels. Scale bar 10 µm. A negative control where DNA was replaced with water is included. C) Plot showing the mean number of BBXs per FOV for labelled CoV (IBV) and the negative controls. Error bars represent the standard deviation of 81 FOVs from one slide. Statistical significance was determined by one-way ANOVA, *$P<0.0001$.

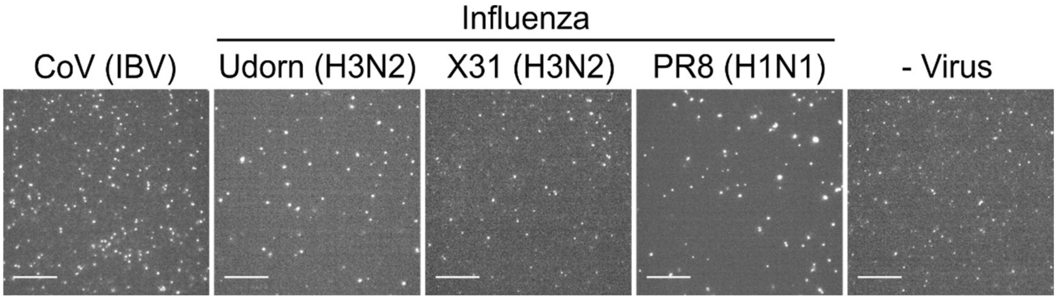


**Sup. Fig. 2. TIRF images of four different viruses.** Representative fields of view (FOVs) of fluorescently labelled CoV (IBV), influenza A (Udorn, X31 and PR8) and a virus-negative control (- Virus). The samples were immobilized and labelled with 0.23 M $SrCl_2$, 1 nM Cy3 (green) DNA and 1 nM Atto647N (red) DNA before being imaged. FOVs from the red channel are shown. Scale bar 10 µm.

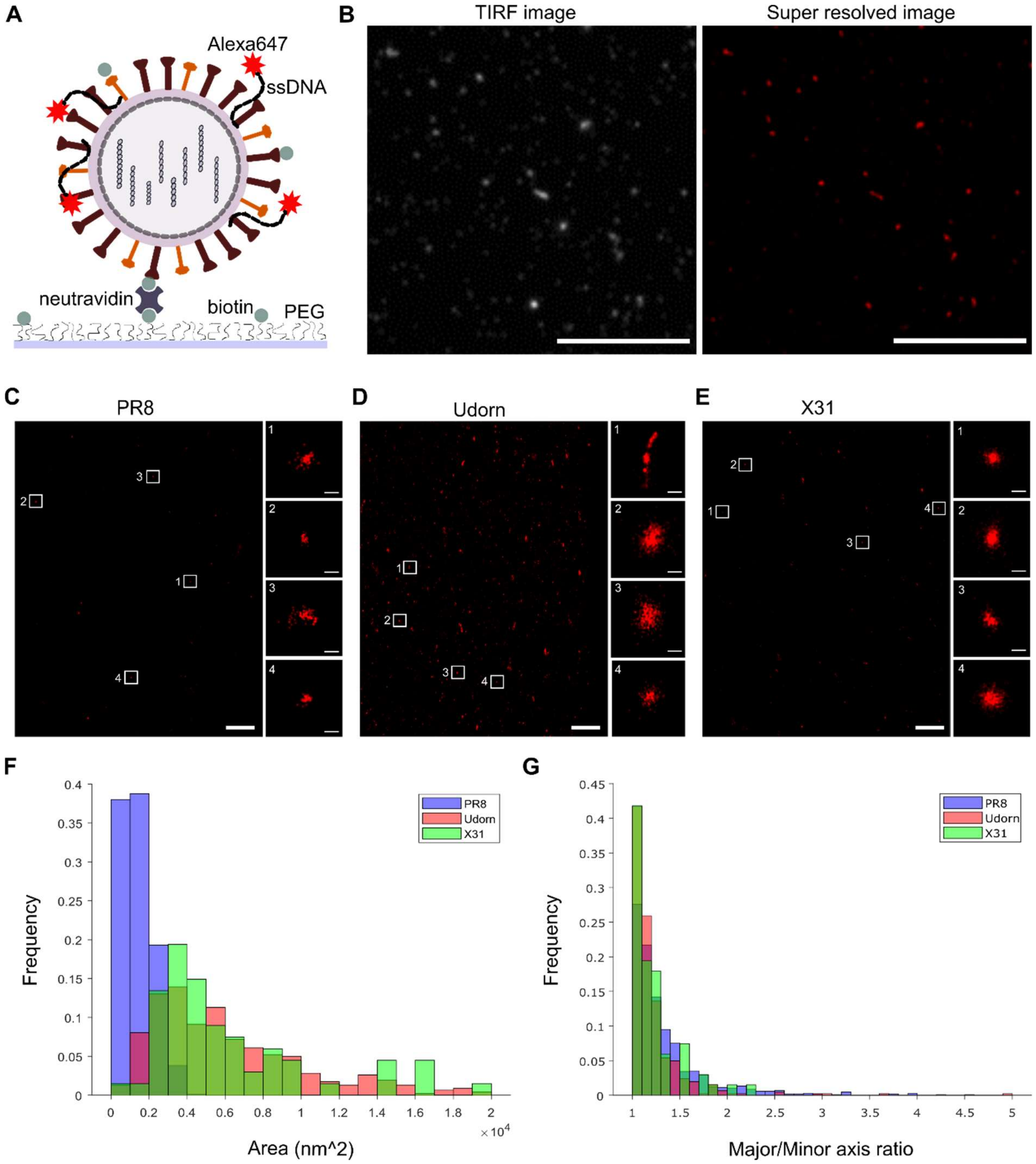


**Sup. Fig. 3 Super resolution images of cation-labelled virus particles.** A) Schematic of immobilisation scheme. Virus particles were biotinylated and subsequently labelled with 0.23 M $CaCl_2$ and 1 nM DNA with a photoswitchable Alexa647 dye. The labelled, biotinylated virus was incubated on a biotin-PEG slide that had been treated with neutravidin, imaging buffer was added and the slide was imaged. B) Representative diffraction-limited TIRF FOV and the corresponding super-resolved image. C) Representative super-resolved image of immobilised PR8 virus. Scale bar 10 µm. White squares highlight zoomed in particles shown to the right, scale bar 100 nm. D) As in C) but for Udorn virus. E) As in C) but for X31 virus. F) Histogram of the areas of PR8, Udorn and X31 (extracted from 4 FOVs of PR8, 2 FOVs of Udorn and 1 FOV of X31). G) Histogram of the major/minor axis ratio of PR8, Udorn and X31.

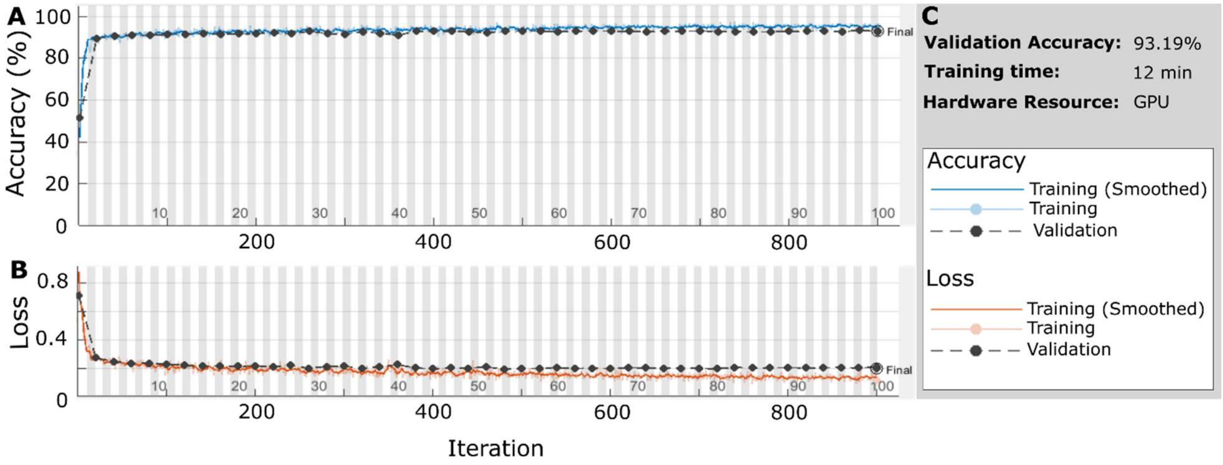


**Sup. Fig. 4 Training Progress for CoV (IBV) and the virus-negative control (- Virus).** A) Graph describing the validation accuracy of the network per iteration. B) Graph showing the loss function value for each iteration. C) Overall validation accuracy and training time.

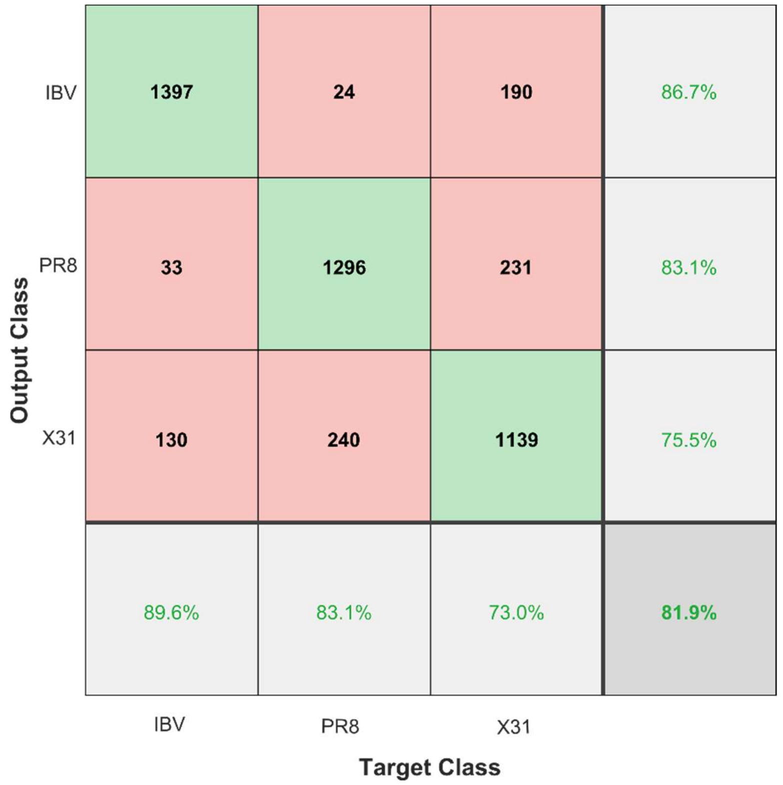


**Sup. Fig. 5. Multi-classifier network validation results for laboratory-grown virus strains.** Confusion matrix showing that a trained network could differentiate between three virus strains (the coronavirus IBV and two influenza strains, PR8 and WSN). 5200 BBXs were obtained for each strain and were then randomly divided into a training dataset (70%) and a validation dataset (30%). The training dataset was used to train the multi-classifier CNN to differentiate IBV from PR8 from WSN. The trained network was validated using the remaining 30% of the data and the results depicted in the confusion matrix. IBV is easily distinguishable from the two influenza strains, which can also be distinguished from each other with high accuracy.

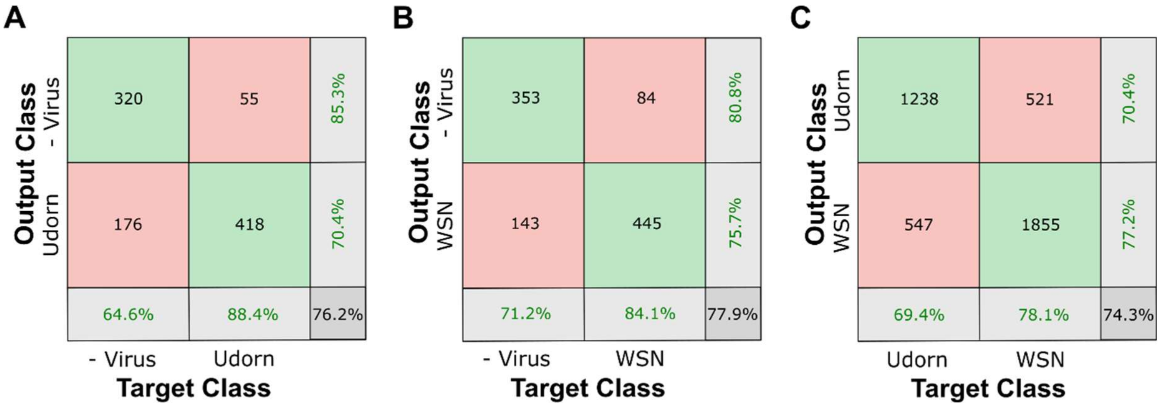


**Sup. Fig. 6. Data taken on a Zeiss Elyra 7 shows similar results.** A) Confusion matrix showing that a trained network could differentiate between Udorn and negative control. B) Confusion matrix showing that a trained network could differentiate between WSN and negative control. C) Confusion matrix showing that a trained network could differentiate between two strains of influenza (WSN and Udorn).

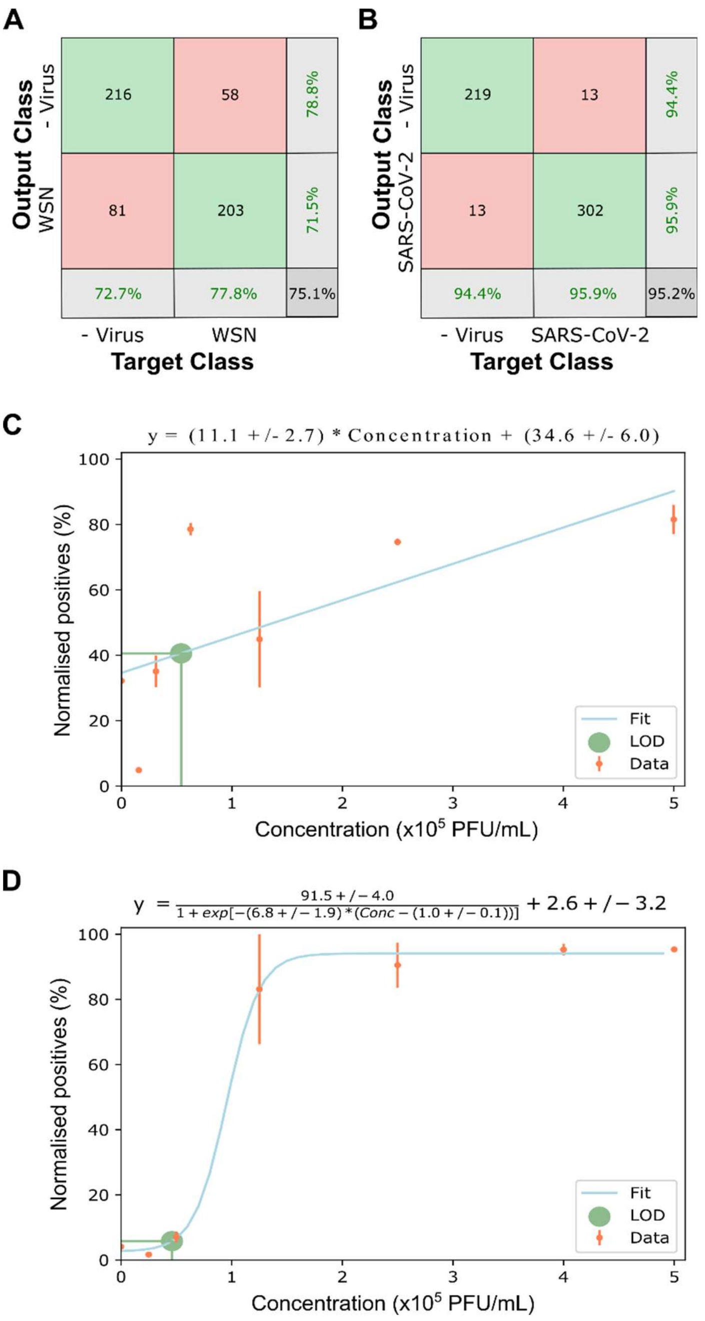


**Sup. Fig. 7. Defining the limit of detection for influenza and SARS-CoV-2.** A) Confusion matrix for a trained model distinguishing influenza strain A/WSN/33 vs. a minus virus control. B) Confusion matrix for SARS-CoV-2 vs. a minus virus control. C) Increasing concentrations of WSN were labelled and imaged, the resulting images were fed into the trained network. The number of normalised positive particles (positive particles/all particles) increased with increasing virus concentration. Error bars represent standard deviation. The limit of detection (LOD) was defined as $5.4 \times 10^4$ PFU/mL. D) As C), for SARS-CoV-2. The limit of detection (LOD) was defined as $4.6 \times 10^4$ PFU/mL.

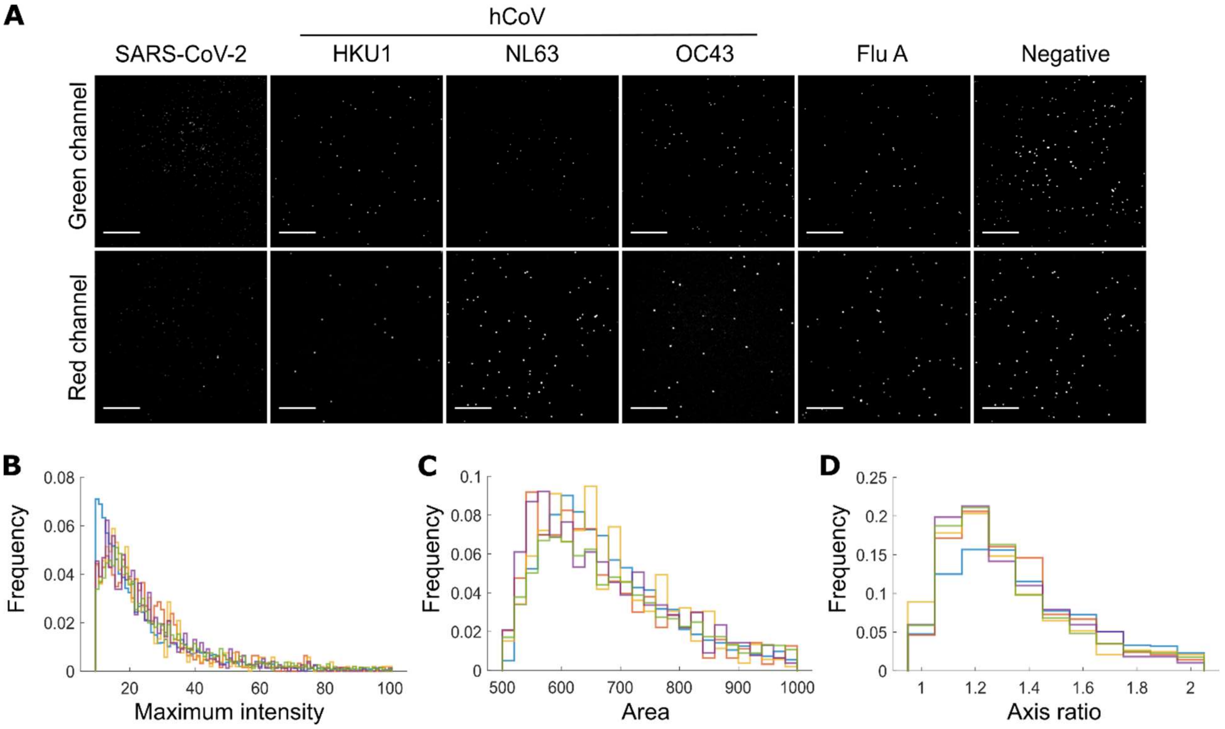


**Sup. Fig. 8. Imaging comparison of clinical samples.** A) Representative FOVs of fluorescently labelled SARS-CoV-2, three seasonal human coronaviruses hCoV, influenza A (Flu A) and a virus-negative control. The samples were immobilized and labelled with 0.23 M $CaCl_2$, 1 nM Cy3 (green) DNA and 1 nM Atto647N (red) DNA before being imaged. Scale bar 10µm. B-D) Normalised frequency plots of the maximum pixel intensity, area and semi-major-to-semi-minor-axis-ratio within the BBXs of five randomly selected SARS-CoV-2 clinical samples to show robustness and reproducibility of labelling. Values taken from 81 FOVs from a single slide for each virus, each sample depicted in a different colour.

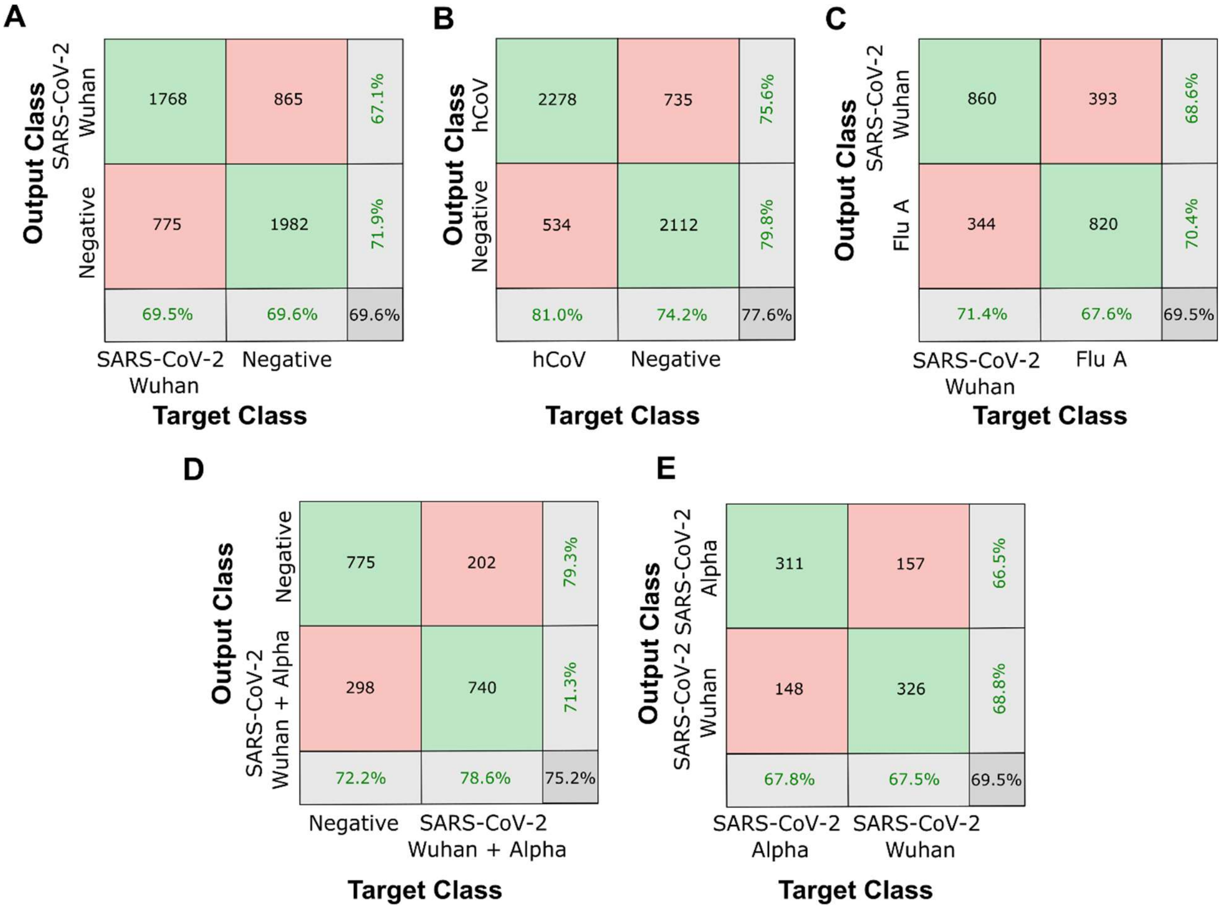


**Sup. Fig. 9. Network validation results for clinical samples.** A) Confusion matrix showing that a trained network could differentiate between SARS-CoV-2 Wuhan strain and negative samples. B) Confusion matrix showing that a trained network could differentiate between seasonal human coronaviruses hCoV and negative samples. C) Confusion matrix showing that a trained network could differentiate between clinical influenza A samples (Flu A) and SARS-CoV-2 Wuhan strain samples. D) Confusion matrix showing that a trained network could differentiate between two strains of SARS-CoV-2 (the original Wuhan strain and the Alpha variant) and negative samples. E) Confusion matrix showing that a trained network could differentiate between clinical samples of two variants of SARS-CoV-2, the original Wuhan strain (SARS-CoV-2 Wuhan) and the Alpha variant.

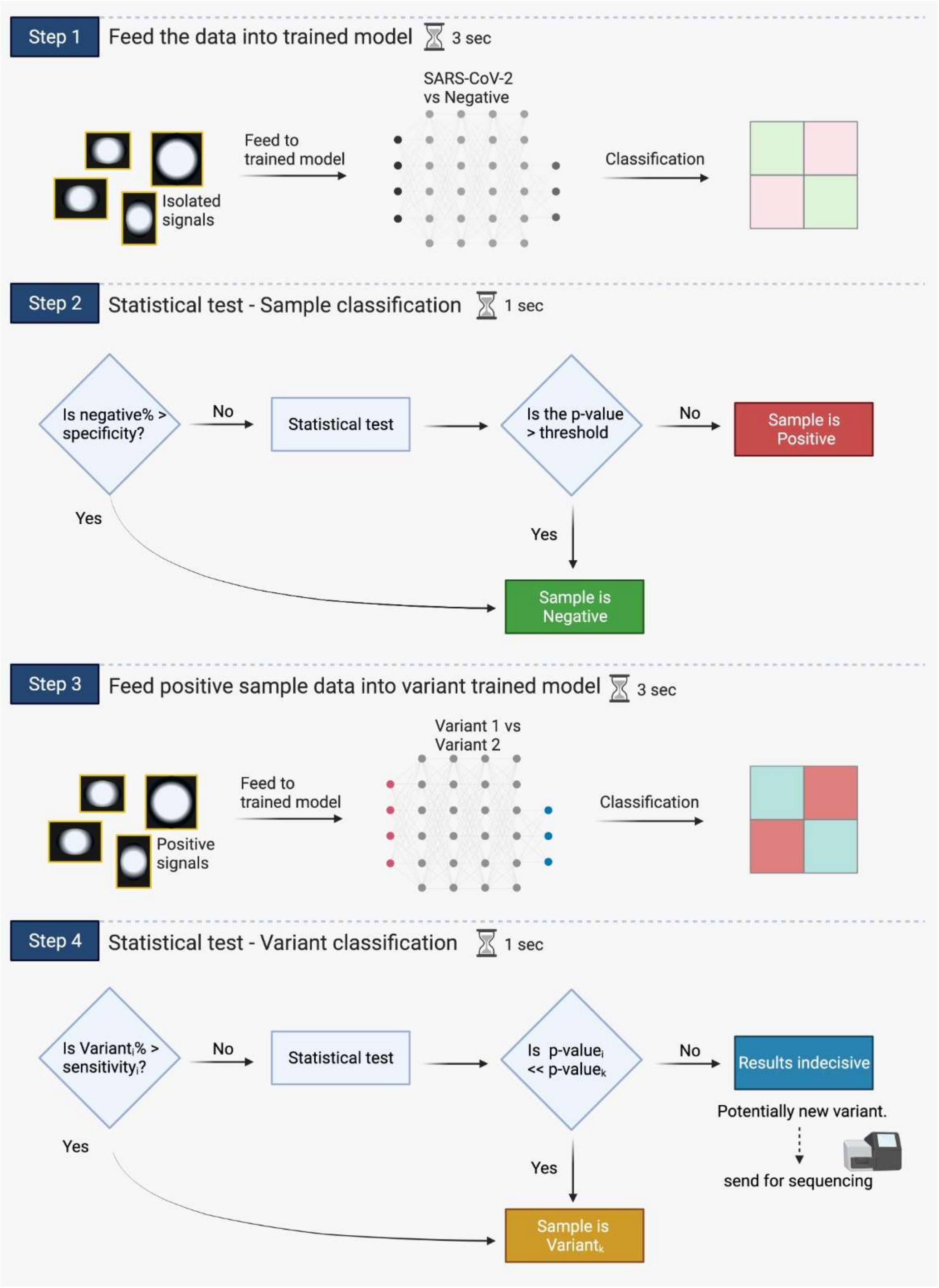


**Sup. Fig. 10. Workflow for sample classification.**

Step 1: Previously unseen samples are imaged, the images are processed into BBXs which are fed through a trained network (in this example, a network that has been trained to distinguish between SARS-CoV-2 and negative samples).

Step 2: Each sample is then classified:

- For each sample, if the percentage of BBXs classified as negative by the trained network (i.e. # negatives/total # of BBXs) is greater than the specificity of the trained model (given in step 3 of the first workflow above), then the sample is automatically classified as a negative.
- If not, then a chi-squared statistical test is performed, where we test the null hypothesis that the sample is negative. For the sample to be classified as positive we need a p-value smaller than our pre-set confidence threshold, in which case we can reject the null hypothesis and classify the sample as positive. If the p-value is greater than our threshold we cannot reject the null hypothesis and therefore the sample is classified as negative.

Step 3: If the sample is classified as SARS-CoV-2, the BBXs identified as positive from that sample can then be passed through a second trained network, in this case to test whether it is Covid-19 variant 1 or Covid-19 variant 2.

Step 4: Variant classification:

- For each sample, if the percentage of BBXs classified as positive by the trained network (i.e. # positives/total # of BBXs) for either variant is greater than the sensitivity for said variant, then the sample is classified as said variant.
- If not, then two chi squared tests are performed, each assuming as their null hypothesis that the sample is one of the two variants. The p-values are then calculated and compared. For a final variant classification, one of the p-values has to be at least 3 orders of magnitude smaller than the other, in which case the sample contains the variant with the bigger p-value. If the two p-values are closer than 3 orders of magnitude, the result is inconclusive, and the sample should be sent for sequencing.

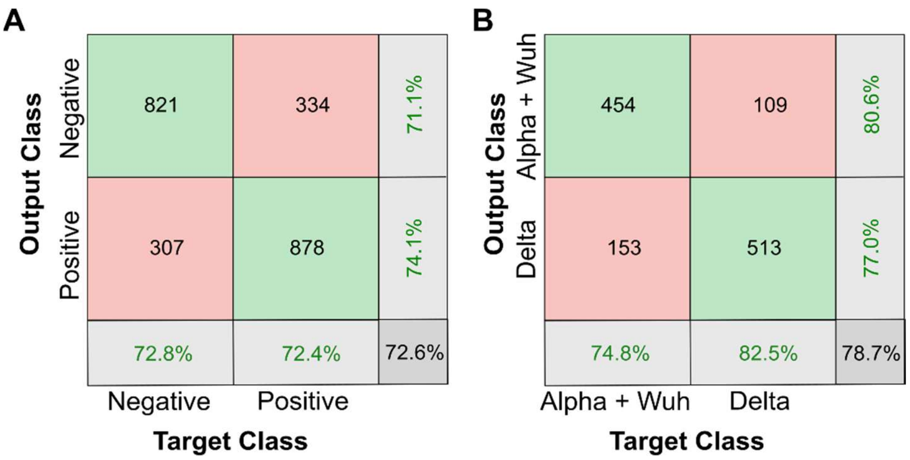


**Sup. Fig. 11. Network validation results for clinical samples.** A) Confusion matrix showing that a trained network could differentiate between SARS-CoV-2 positive and negative samples. B) Confusion matrix showing that a trained network could differentiate between the Delta SARS-CoV-2 variant and a combination of the Wuhan and Alpha variant strains of SARS-CoV-2.

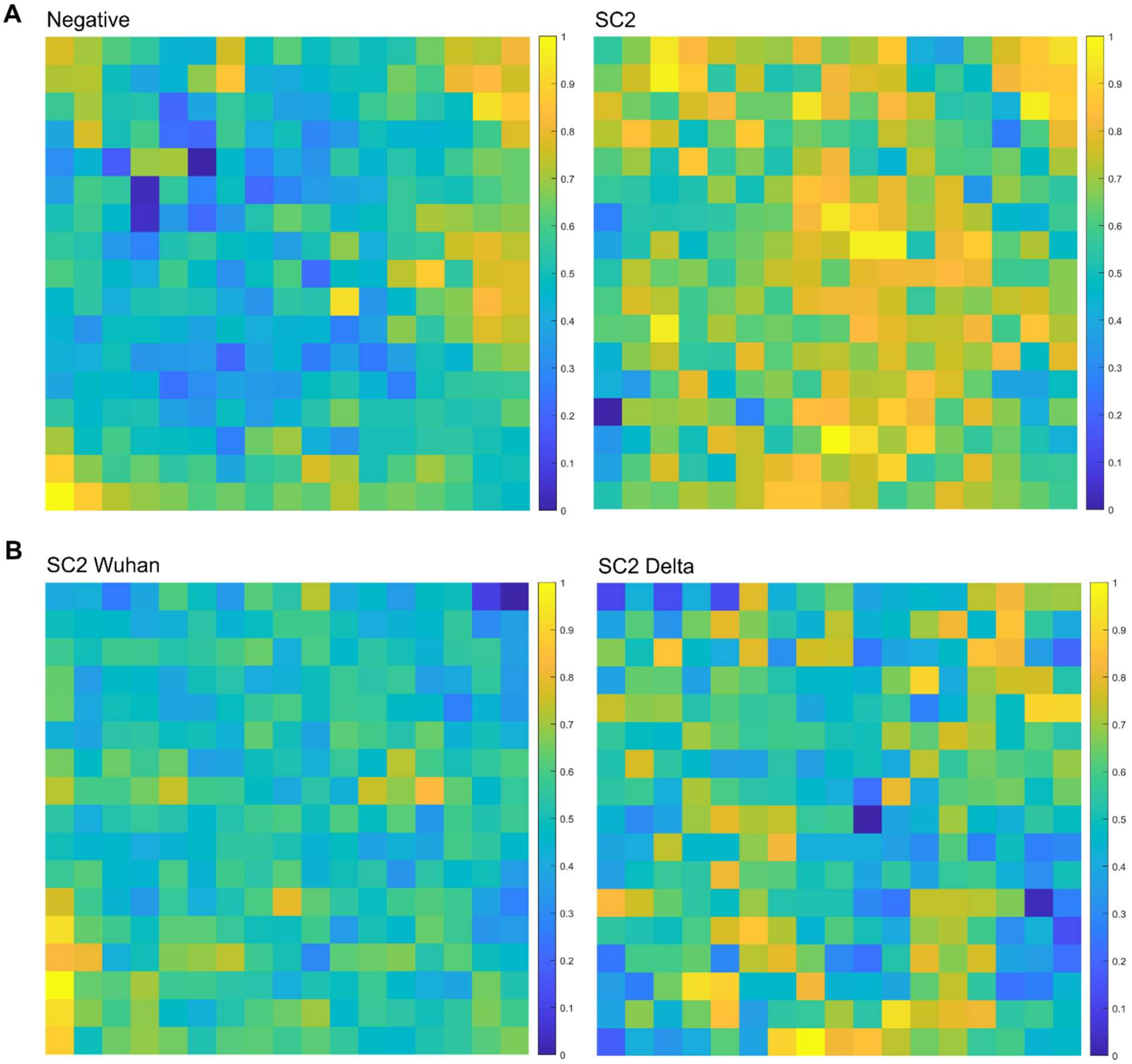


**Sup. Fig. 12. Learned pattern visualisation by the convolutional neural network using the Matlab DeepDreamImage function.** A) Patterns ‘seen’ by a network trained to differentiate between negative (left panel) and SARS-CoV-2 positive (SC2; right panel) samples. An image was forwarded through the network, and the gradient of the image with respect to the activations of a particular layer was calculated (in this case the fully connected layer). The image was modified to maximise these activations, enhancing the patterns 'seen' by the network. B) Patterns ‘seen’ by a network trained to differentiate between the original Wuhan variant strain of SARS-CoV-2 (SC2 Wuhan; left panel) and the Delta SARS-CoV-2 variant (SC2 Delta; right panel).

**Table 1. Data on clinical samples used for training and validating the network.** Multiple samples were imaged over three different days (described as -1, -2, -3 etc.). 70% of the BBXs generated over the three days were randomly selected for network training, while 30% were reserved for network validation (results depicted in the confusion matrices). Abbreviations used: hCoV – seasonal human coronaviruses (including the NL63, OC43 and HKU1 types), Neg – negative, SC2 – SARS-CoV-2 (original Wuhan variant), Flu A – Influenza A. 'Microscope 1' was a Nanoimager equipped with a Hamamatsu Flash 4 LT.1 sCMOS camera and 'Microscopes 2 and 3' were Nanoimagers equipped with a Hamamatsu Flash4 V3 sCMOS camera; in all other respects, the systems were identical.

| **Network** | **hCoV vs Negative** (Microscope 1) | | | | | |
|---|---|---|---|---|---|---|
| | **Identifier** | **RT-PCR** | **Total # of BBXs** | **Identifier** | **RT-PCR** | **Total # of BBXs** |
| | T-Neg-1 | Neg | 1507 | T-hCov-3-3 | HKU1 | 148 |
| | T-Neg-2 | Neg | 562 | T-hCov-4 | NL63 | 377 |
| | T-Neg-3 | Neg | 451 | T-hCov-4-1 | NL63 | 80 |
| | T-Neg-4 | Neg | 378 | T-hCov-4-2 | NL63 | 279 |
| | T-Neg-1-1 | Neg | 5173 | T-hCov-5 | NL63 | 500 |
| | T-Neg-2-1 | Neg | 122 | T-hCov-5-1 | NL63 | 513 |
| | T-Neg-3-1 | Neg | 568 | T-hCov-5-2 | NL63 | 823 |
| | T-Neg-4-1 | Neg | 158 | T-hCov-6 | NL63 | 453 |
| | T-Neg-4-2 | Neg | 571 | T-hCov-6-1 | NL63 | 207 |
| | T-hCov-1 | HKU1 | 229 | T-hCov-6-2 | NL63 | 292 |
| | T-hCov-1-1 | HKU1 | 290 | T-hCov-7 | OC43 | 268 |
| | T-hCov-1-2 | HKU1 | 104 | T-hCov-7-1 | OC43 | 131 |
| | T-hCov-1-3 | HKU1 | 203 | T-hCov-7-2 | OC43 | 254 |
| | T-hCov-2 | HKU1 | 169 | T-hCov-8 | OC43 | 897 |
| | T-hCov-2-1 | HKU1 | 92 | T-hCov-8-1 | OC43 | 569 |
| | T-hCov-2-2 | HKU1 | 136 | T-hCov-8-2 | OC43 | 342 |
| | T-hCov-3 | HKU1 | 330 | T-hCov-9 | OC43 | 500 |
| | T-hCov-3-1 | HKU1 | 918 | T-hCov-9-1 | OC43 | 145 |
| | T-hCov-3-2 | HKU1 | 129 | | | |
| | | | | | **Total # of BBXs in dataset** | 18863 |
| | **SARS-CoV-2 Wuhan vs hCoV** (Microscope 1) | | | | | |
| | **Identifier** | **RT-PCR** | **Total # of BBXs** | **Identifier** | **RT-PCR** | **Total # of BBXs** |
| | T-SC2-1 | SC2 | 696 | T-hCov-3-2 | HKU1 | 129 |
| | T-SC2-1-1 | SC2 | 3320 | T-hCov-3-3 | HKU1 | 148 |
| | T-SC-2 | SC2 | 370 | T-hCov-4 | NL63 | 377 |
| | T-SC-2-1 | SC2 | 223 | T-hCov-4-1 | NL63 | 80 |
| | T-SC-2-2 | SC2 | 131 | T-hCov-4-2 | NL63 | 279 |
| | T-SC-3 | SC2 | 662 | T-hCov-5 | NL63 | 500 |
| | T-SC-3-1 | SC2 | 214 | T-hCov-5-1 | NL63 | 513 |
| | T-SC-4 | SC2 | 861 | T-hCov-5-2 | NL63 | 823 |
| | T-SC-4-1 | SC2 | 218 | T-hCov-6 | NL63 | 453 |
| | T-SC-4-2 | SC2 | 1783 | T-hCov-6-1 | NL63 | 207 |
| | T-hCov-1 | HKU1 | 229 | T-hCov-6-2 | NL63 | 292 |
| | T-hCov-1-1 | HKU1 | 290 | T-hCov-7 | OC43 | 268 |
| | T-hCov-1-2 | HKU1 | 104 | T-hCov-7-1 | OC43 | 131 |
| | T-hCov-1-3 | HKU1 | 203 | T-hCov-7-2 | OC43 | 254 |
| | T-hCov-2 | HKU1 | 169 | T-hCov-8 | OC43 | 897 |
| | T-hCov-2-1 | HKU1 | 92 | T-hCov-8-1 | OC43 | 569 |
| | T-hCov-2-2 | HKU1 | 136 | T-hCov-8-2 | OC43 | 342 |
| | T-hCov-3 | HKU1 | 330 | T-hCov-9 | OC43 | 500 |
| | T-hCov-3-1 | HKU1 | 918 | T-hCov-9-1 | OC43 | 145 |
| | | | | | **Total # of BBXs in dataset** | 17851 |
| | **SARS-CoV-2 Wuhan vs Negative** (Microscope 1) | | | | | |
| | **Identifier** | **RT-PCR** | **Total # of BBXs** | **Identifier** | **RT-PCR** | **Total # of BBXs** |
| | T-SC2-1 | SC2 | 696 | T-SC-4-2 | SC2 | 1783 |
| | T-SC2-1-1 | SC2 | 3320 | T-Neg-1 | Neg | 1507 |
| | T-SC-2 | SC2 | 370 | T-Neg-2 | Neg | 562 |
| | T-SC-2-1 | SC2 | 223 | T-Neg-3 | Neg | 451 |

| | | | | | | |
|---|---|---|---|---|---|---|
| | T-SC-2-2 | SC2 | 131 | T-Neg-4 | Neg | 378 |
| | T-SC-3 | SC2 | 662 | T-Neg-1-1 | Neg | 5173 |
| | T-SC-3-1 | SC2 | 214 | T-Neg-2-1 | Neg | 122 |
| | T-SC-4 | SC2 | 861 | T-Neg-3-1 | Neg | 568 |
| | T-SC-4-1 | SC2 | 218 | T-Neg-4-1 | Neg | 729 |
| | | | | | **Total # of BBXs in dataset** | 17968 |
| | **Flu A vs Negative** (Microscope 1) | | | | | |
| | **Identifier** | **RT-PCR** | **Total # of BBXs** | **Identifier** | **RT-PCR** | **Total # of BBXs** |
| | T-Neg-5 | Neg | 514 | T-Neg-4 | Neg | 378 |
| | T-Neg-5-1 | Neg | 337 | T-Neg-4-1 | Neg | 158 |
| | T-Neg-5-2 | Neg | 471 | T-Neg-4-2 | Neg | 571 |
| | T-Neg-2 | Neg | 652 | T-Neg-4-3 | Neg | 154 |
| | T-Neg-2-1 | Neg | 77 | T-Neg-4-4 | Neg | 233 |
| | T-Neg-2-2 | Neg | 45 | T-FluA-1 | Flu A | 1782 |
| | T-Neg-2-3 | Neg | 242 | T-FluA-1-1 | Flu A | 289 |
| | T-Neg-3 | Neg | 451 | T-FluA-1-2 | Flu A | 246 |
| | T-Neg-3-1 | Neg | 568 | T-FluA-2 | Flu A | 61 |
| | T-Neg-3-2 | Neg | 398 | T-FluA-2-1 | Flu A | 658 |
| | T-Neg-3-3 | Neg | 150 | T-FluA-3 | Flu A | 1177 |
| | T-Neg-3-4 | Neg | 374 | | | |
| | | | | | **Total # of BBXs in dataset** | 10461 |
| | **SARS-COV-2 Wuhan vs Flu A** (Microscope 1) | | | | | |
| | **Identifier** | **RT-PCR** | **Total # of BBXs** | **Identifier** | **RT-PCR** | **Total # of BBXs** |
| | T-FluA-5 | Flu A | 289 | T-SC-2-1 | SC2 | **223** |
| | T-FluA-5-1 | Flu A | 246 | T-SC-2-3 | SC2 | 318 |
| | T-FluA-5-2 | Flu A | 2416 | T-SC-4-3 | SC2 | 122 |
| | T-FluA-6 | Flu A | 188 | T-SC-4-4 | SC2 | 200 |
| | T-FluA-6-1 | Flu A | 748 | T-SC-4-5 | SC2 | 546 |
| | T-FluA-6-2 | Flu A | 158 | T-SC-5 | SC2 | 170 |
| | T-SC2-1-2 | SC2 | 248 | T-SC-5-1 | SC2 | 260 |
| | T-SC2-1-3 | SC2 | 209 | T-SC-5-2 | SC2 | 630 |
| | T-SC2-1-4 | SC2 | 236 | T-SC-5-3 | SC2 | 823 |
| | | | | | **Total # of BBXs in dataset** | 8060 |
| | **SARS-CoV-2 Alpha vs SARS-CoV-2 Wuhan** (Microscope 2) | | | | | |
| | **Identifier** | **RT-PCR** | **Total # of BBXs** | **Identifier** | **RT-PCR** | **Total # of BBXs** |
| | T-SC2-6 | B.1.1.7 | 631 | T-SC2-9 | SC2 | 483 |
| | T-SC2-7 | B.1.1.7 | 663 | T-SC2-10 | SC2 | 511 |
| | T-SC2-8 | B.1.1.7 | 235 | T-SC2-11 | SC2 | 616 |
| | | | | | **Total # of BBXs in dataset** | 3139 |
| | **SARS-CoV-2 (Wuhan and Alpha) vs Negative** (Microscope 2) | | | | | |
| | **Identifier** | **RT-PCR** | **Total # of BBXs** | **Identifier** | **RT-PCR** | **Total # of BBXs** |
| | T-SC2-6 | B.1.17 | 631 | T-SC2-11 | SC2 | 616 |
| | T-SC2-12 | B.1.1.7 | 127 | T-Neg-5 | Neg | 655 |
| | T-SC2-13 | B.1.1.7 | 771 | T-Neg-6 | Neg | 1318 |
| | T-SC2-9 | SC2 | 483 | T-Neg-7 | Neg | 595 |
| | T-SC2-10 | SC2 | 511 | T-Neg-8 | Neg | 1008 |
| | | | | | **Total # of BBXs in dataset** | 6715 |
| | **SARS-CoV-2 Alpha vs Negative** (Microscope 2) | | | | | |
| | **Identifier** | **RT-PCR** | **Total # of BBXs** | **Identifier** | **RT-PCR** | **Total # of BBXs** |
| | T-SC-2-6 | B.1.1.7 | 631 | T-Neg-5 | Neg | 655 |
| | T-SC-2-7 | B.1.1.7 | 663 | T-Neg-7 | Neg | 469 |
| | T-SC-2-8 | B.1.1.7 | 235 | T-Neg-9 | Neg | 595 |
| | | | | | **Total # of BBXs in dataset** | 3248 |
| | **SARS-CoV-2 vs Negative** (Microscope 3) | | | | | |
| | **Identifier** | **RT-PCR** | **Total # of BBXs** | **Identifier** | **RT-PCR** | **Total # of BBXs** |
| | NEG-J-2 | Negative | 792 | NEG-J-30 | Negative | 310 |
| | NEG-J-3 | Negative | 135 | SC2-WT-02 | SARS-CoV-2 | 483 |
| | NEG-J-4 | Negative | 182 | SC2-WT-07 | SARS-CoV-2 | 511 |
| | NEG-J-5 | Negative | 451 | SC2-WT-13 | SARS-CoV-2 | 616 |
| | NEG-J-6 | Negative | 672 | SC2-D-2 | SARS-CoV-2 | 500 |
| | NEG-J-12 | Negative | 1218 | SC2 lab grown | N/A | 582 |
| | | | | | **Total # of BBXs in dataset** | 6452 |
| | **Wuhan + Alpha vs Delta variant** (Microscope 3) | | | | | |

| | Identifier | RT-PCR | Total # of BBXs | Identifier | RT-PCR | Total # of BBXs |
|---|---|---|---|---|---|---|
| | SC2-A-01 | SARS-CoV-2 | 360 | SC2-D-3 | SARS-CoV-2 | 31 |
| | SC2-A-03 | SARS-CoV-2 | 188 | SC2-D-4 | SARS-CoV-2 | 9 |
| | SC2-A-09 | SARS-CoV-2 | 1022 | SC2-D-9 | SARS-CoV-2 | 266 |
| | SC2-WT-04 | SARS-CoV-2 | 491 | SC2-D-10 | SARS-CoV-2 | 81 |
| | SC2-WT-05 | SARS-CoV-2 | 572 | SC2-D-12 | SARS-CoV-2 | 455 |
| | SC2-WT-11 | SARS-CoV-2 | 460 | SC2-D-13 | SARS-CoV-2 | 236 |
| | SC2-D-1 | SARS-CoV-2 | 1680 | SC2-D-29 | SARS-CoV-2 | 536 |
| | | | | | **Total # of BBXs in dataset** | 6387 |

**Table 2. Results of independent testing of the network on samples not seen before (51 samples).** Abbreviations used: hCoV – seasonal human coronaviruses (including the NL63, OC43 and HKU1 types), Neg – negative, na – statistical test not needed as the percentage of BBXs classified as negative by the trained network (i.e. # negatives/total # of BBXs) is greater than the specificity of the trained model, SC2 – SARS-CoV-2, Flu A – Influenza A, SC2-S0 – SARS-CoV-2 spike gene target failure in RT-PCR indicative of the Alpha variant. Incorrectly classified sample highlighted in turquoise.

| **Network** | **Identifier** | **RT-PCR** | | **Total # of BBXs** | | **P-value** | **Result** |
|---|---|---|---|---|---|---|---|
| 'Microscope 1' | | | | hCov | Negative | Threshold: 0.01 | |
| **hCoV vs Negative** | hCoV-1 | NL63 | 6467 | 4271 | 2196 | 5.18E-10 | hCoV |
| | hCoV-2 | NL63 | 2672 | 1307 | 1365 | 5.184-21 | hCoV |
| | hCoV-3 | NL63 | 227 | 111 | 116 | 0.006 | hCoV |
| | hCoV-4 | OC43 | 2239 | 1706 | 533 | 9.25E-268 | hCoV |
| | hCoV-5 | OC43 | 653 | 442 | 211 | 2.80E-47 | hCoV |
| | hCoV-6 | OC43 | 840 | 555 | 285 | 1.13E-53 | hCoV |
| | hCoV-7 | HKU1 | 1342 | 1087 | 255 | 2.09E-206 | hCoV |
| | hCoV-8 | HKU1 | 1898 | 1069 | 829 | 9.69E-48 | hCoV |
| | hCoV-9 | HKU1 | 6745 | 3797 | 2948 | 2.82E-164 | hCoV |
| | NEG-1 | Neg | 514 | 198 | 316 | 0.49 | Neg |
| | NEG-2 | Neg | 398 | 141 | 257 | na | Neg |
| | NEG-3 | Neg | 154 | 29 | 125 | na | Neg |
| 'Microscope 1' | | | | SC2 | hCov | Threshold: 0.01 | |
| **SARS-CoV-2 Wuhan vs hCoV** | SC2-1 | SARS-CoV-2 | 5589 | 3653 | 1936 | 5.18E-10 | SC2 |
| | SC2-2 | SARS-CoV-2 | 1762 | 1312 | 450 | 5.14E-21 | SC2 |
| | SC2-3 | SARS-CoV-2 | 284 | 86 | 198 | 0.917 | hCoV |
| | hCoV-7 | HKU1 | 1342 | 159 | 1183 | na | hCoV |
| | hCoV-8 | HKU1 | 1898 | 593 | 1305 | 0.237 | hCoV |
| | hCoV-10 | HKU1 | 3182 | 654 | 2528 | na | hCoV |
| | hCoV-11 | OC43 | 264 | 8 | 256 | na | hCoV |
| | hCoV-12 | OC43 | 479 | 21 | 458 | na | hCoV |
| | hCoV-13 | OC43 | 7966 | 2245 | 5721 | na | hCoV |
| | hCoV-14 | NL63 | 305 | 12 | 293 | na | hCoV |
| | hCoV-15 | NL63 | 968 | 89 | 879 | na | hCoV |
| | hCoV-16 | NL63 | 2269 | 349 | 1920 | na | hCoV |
| 'Microscope 1' | | | | SC2 | Neg | Threshold: 0.01 | |
| **SARS-CoV-2 Wuhan vs Negative** | SC2-4 | SARS-CoV-2 | 200 | 150 | 50 | 7.93E-21 | SC2 |
| | SC2-5 | SARS-CoV-2 | 560 | 503 | 57 | 1.07E-114 | SC2 |
| | SC2-6 | SARS-CoV-2 | 529 | 496 | 33 | 7.58E-127 | SC2 |
| | SC2-7 | SARS-CoV-2 | 173 | 148 | 25 | 1.11E-30 | SC2 |
| | SC2-8 | SARS-CoV-2 | 150 | 135 | 15 | 2.89E-32 | SC2 |
| | SC2-9 | SARS-CoV-2 | 8095 | 3575 | 4520 | 6.91E-4 | SC2 |
| | Neg-4 | Neg | 158 | 28 | 130 | na | Neg |
| | Neg-5 | Neg | 568 | 240 | 328 | 0.982 | Neg |
| | Neg-6 | Neg | 77 | 16 | 61 | na | Neg |
| | Neg-7 | Neg | 337 | 132 | 215 | na | Neg |
| 'Microscope 1' | | | | Flu A | Neg | Threshold: 0.01 | |
| **Flu A vs Negative** | Flu A -1 | Flu A | 311 | 263 | 48 | 2.08E-08 | Flu A |
| | Flu A -2 | Flu A | 195 | 163 | 32 | 3.46E-05 | Flu A |
| | Flu A -3 | Flu A | 24383 | 24231 | 152 | 0.00 | Flu A |
| | Flu A -4 | Flu A | 212 | 185 | 27 | 4.13E-08 | Flu A |
| | Neg-8 | Neg | 1507 | 41 | 1466 | na | Neg |
| | Neg-9 | Neg | 374 | 259 | 115 | 0.22 | Neg |
| | Neg-10 | Neg | 354 | 235 | 119 | 0.14 | Neg |
| 'Microscope 1' | | | | SC2 | Flu A | Threshold: 0.01 | |
| **SARS-CoV-2 Wuhan vs Flu A** | SC2-10 | SARS-CoV-2 | 1783 | 1551 | 232 | 0.00 | SC2 |
| | SC2-11 | SARS-CoV-2 | 1340 | 818 | 522 | 2.54E-81 | SC2 |
| | SC2-12 | SARS-CoV-2 | 13915 | 5413 | 8502 | 3.40E-21 | SC2 |
| | Flu A -1 | Flu A | 311 | 106 | 205 | na | Flu A |

| | | | | | | | | |
|---|---|---|---|---|---|---|---|---|
| | Flu A -2 | Flu A | 195 | 53 | 142 | 0.270 | | Flu A |
| | Flu A -3 | Flu A | 24383 | 2871 | 21512 | na | | Flu A |
| | Flu A -4 | Flu A | 212 | 51 | 161 | na | | Flu A |
| ‘Microscope 2’ | | | | B.1.1.7 | Negative | Threshold: 0.01 | | |
| **SARS-CoV-2 Alpha vs Negative** | SC2-13 | SC2-S0 | 4979 | 2538 | 2441 | 0.00 | | B.1.1.7 |
| | SC2-14 | SC2-S0 | 631 | 602 | 29 | 0.00 | | B.1.1.7 |
| | SC2-15 | SC2-S0 | 127 | 117 | 10 | 2.18E-61 | | B.1.1.7 |
| | SC2-16 | SC2-S0 | 771 | 738 | 33 | 0.00 | | B.1.1.7 |
| | Neg-8 | Neg | 4166 | 22 | 4144 | na | | Neg |
| | Neg-9 | Neg | 2888 | 130 | 2758 | na | | Neg |
| | Neg-10 | Neg | 3369 | 947 | 2422 | 0.147 | | Neg |
| ‘Microscope 2’ | | | | SC2 | Negative | Threshold: 0.01 | | |
| **SARS-CoV-2 (Wuhan + Alpha) vs Negative** | SC2-17 | SC2-S0 | 5024 | 2309 | 2715 | 0.000 | | SC2 |
| | SC2-14 | SC2-S0 | 631 | 563 | 68 | 0.000 | | SC2 |
| | SC2-15 | SC2-S0 | 127 | 97 | 30 | 0.000 | | SC2 |
| | SC2-16 | SC2-S0 | 771 | 709 | 62 | 0.000 | | SC2 |
| | SC2-18 | SC2-S0 | 2492 | 2254 | 238 | 0.000 | | SC2 |
| | SC2-19 | SC2 | 4058 | 1302 | 2756 | 0.004 | | SC2 |
| | SC2-20 | SC2 | 1609 | 1410 | 199 | 0.000 | | SC2 |
| | Neg-8 | Neg | 4166 | 33 | 4133 | na | | Neg |
| | Neg-9 | Neg | 2888 | 164 | 2724 | na | | Neg |
| | Neg-10 | Neg | 2422 | 32 | 2390 | na | | Neg |
| | Neg-11 | Neg | 2435 | 741 | 1694 | 0.64 | | Neg |
| ‘Microscope 2’ | | | | SC2 | B.1.1.7 | p-SC2 | p-B.1.1.7 | |
| **SARS-CoV-2 Alpha vs SARS-CoV-2 Wuhan** | SC2-17 | SC2-S0 | 2309 | 981 | 1328 | 1.683E-138 | 3.20E-22 | B.1.1.7 |
| | SC2-14 | SC2-S0 | 563 | 116 | 447 | 3.21E-121 | 4.06E-13 | B.1.1.7 |
| | SC2-15 | SC2-S0 | 97 | 33 | 64 | 4.92E-12 | 0.831 | B.1.1.7 |
| | SC2-16 | SC2-S0 | 709 | 155 | 554 | 4.17E-144 | 2.84E-10 | B.1.1.7 |
| | SC2-18 | SC2-S0 | 2254 | 1051 | 1203 | 5.21E-94 | 4.43E-43 | B.1.1.7 |
| | SC2-19 | SC2 | 1302 | 725 | 577 | 3.82E-18 | 7.31E-68 | SC2 |
| | SC2-20 | SC2 | 1410 | 776 | 634 | 1.24E-21 | 2.60E-69 | SC2 |

**Table 3. Results of independent testing of the network on samples not seen before (104 samples).** Abbreviations used: Neg – negative, NA – statistical test not needed as the percentage of BBXs classified as negative by the trained network (i.e. # negatives/total # of BBXs) is greater than the specificity of the trained model, SC2 – SARS-CoV-2. Samples with less than the 5 BBXs needed for the chi-squared test are highlighted in yellow and incorrectly classified samples are highlighted in turquoise.

| **Network** | **Identifier** | **RT-PCR** | **# of BBXs** | | | **P-value** | **Result** |
|---|---|---|---|---|---|---|---|
| 'Microscope 3' | | | Total | SC2 | Negative | Threshold: 0.012 | |
| **SARS-CoV-2 vs Negative** | **Neg-2-01** | **Negative** | **41** | **1** | **40** | **NA** | **NEGATIVE** |
| | Neg-2-02 | Negative | 434 | 71 | 363 | NA | NEGATIVE |
| | Neg-2-03 | Negative | 820 | 154 | 666 | NA | NEGATIVE |
| | Neg-2-04 | Negative | 1126 | 228 | 898 | NA | NEGATIVE |
| | Neg-2-05 | Negative | 716 | 135 | 581 | NA | NEGATIVE |
| | Neg-2-06 | Negative | 1390 | 363 | 1027 | NA | NEGATIVE |
| | Neg-2-07 | Negative | 891 | 250 | 641 | NA | NEGATIVE |
| | Neg-2-08 | Negative | 1763 | 491 | 1272 | NA | NEGATIVE |
| | Neg-2-09 | Negative | 235 | 33 | 202 | NA | NEGATIVE |
| | Neg-J-02 | Negative | 922 | 145 | 777 | NA | NEGATIVE |
| | Neg-J-03 | Negative | 274 | 42 | 232 | NA | NEGATIVE |
| | Neg-J-04 | Negative | 182 | 31 | 151 | NA | NEGATIVE |
| | Neg-J-05 | Negative | 1078 | 139 | 939 | NA | NEGATIVE |
| | Neg-J-06 | Negative | 932 | 124 | 808 | NA | NEGATIVE |
| | Neg-J-07 | Negative | 1383 | 181 | 1202 | NA | NEGATIVE |
| | Neg-J-08 | Negative | 571 | 117 | 454 | NA | NEGATIVE |
| | Neg-J-09 | Negative | 768 | 116 | 652 | NA | NEGATIVE |
| | Neg-J-10 | Negative | 759 | 97 | 662 | NA | NEGATIVE |
| | Neg-J-11 | Negative | 154 | 29 | 125 | NA | NEGATIVE |
| | NEG-J-12 | Negative | 1341 | 428 | 913 | 0.468048338 | NEGATIVE |
| | Neg-J-13 | Negative | 988 | 294 | 694 | NA | NEGATIVE |
| | Neg-J-14 | Negative | 4 | 3 | 1 | INCONCLUSIVE | INCONCLUSIVE |
| | Neg-J-15 | Negative | 897 | 286 | 611 | 0.566986783 | NEGATIVE |
| | Neg-J-16 | Negative | 248 | 48 | 200 | NA | NEGATIVE |
| | Neg-J-17 | Negative | 65 | 14 | 51 | NA | NEGATIVE |
| | Neg-J-18 | Negative | 156 | 14 | 142 | NA | NEGATIVE |
| | Neg-J-19 | Negative | 216 | 31 | 185 | NA | NEGATIVE |
| | Neg-J-20 | Negative | 1163 | 392 | 771 | 0.04601353 | NEGATIVE |
| | Neg-J-21 | Negative | 634 | 144 | 490 | NA | NEGATIVE |
| | Neg-J-22 | Negative | 53 | 13 | 40 | NA | NEGATIVE |
| | Neg-J-23 | Negative | 218 | 33 | 185 | NA | NEGATIVE |
| | Neg-J-24 | Negative | 580 | 155 | 425 | NA | NEGATIVE |
| | Neg-J-25 | Negative | 456 | 121 | 335 | NA | NEGATIVE |
| | Neg-J-26 | Negative | 567 | 135 | 432 | NA | NEGATIVE |
| | Neg-J-27 | Negative | 377 | 72 | 305 | NA | NEGATIVE |
| | Neg-J-28 | Negative | 243 | 57 | 186 | NA | NEGATIVE |
| | Neg-J-29 | Negative | 1257 | 508 | 749 | 5.34E-13 | POSITIVE |
| | Neg-J-30 | Negative | 310 | 44 | 266 | NA | NEGATIVE |
| | Neg-J-31 | Negative | 46 | 7 | 39 | NA | NEGATIVE |
| | Neg-J-32 | Negative | 119 | 14 | 105 | NA | NEGATIVE |
| | Neg-J-33 | Negative | 316 | 75 | 241 | NA | NEGATIVE |
| | Neg-J-34 | Negative | 1593 | 445 | 1148 | NA | NEGATIVE |
| | Neg-J-36 | Negative | 973 | 185 | 788 | NA | NEGATIVE |
| | Neg-J-37 | Negative | 46 | 6 | 40 | NA | NEGATIVE |
| | Neg-J-38 | Negative | 521 | 71 | 450 | NA | NEGATIVE |
| | Neg-J-39 | Negative | 1495 | 357 | 1138 | NA | NEGATIVE |
| | Neg-J-40 | Negative | 1026 | 249 | 777 | NA | NEGATIVE |
| | Neg-J-41 | Negative | 73 | 11 | 62 | NA | NEGATIVE |
| | Neg-J-42 | Negative | 985 | 304 | 681 | NA | NEGATIVE |
| | NEG-J-43 | Negative | 1 | 0 | 1 | INCONCLUSIVE | INCONCLUSIVE |
| | NEG-J-44 | Negative | 14 | 2 | 12 | NA | NEGATIVE |

| | | | | | | | |
|---|---|---|---|---|---|---|---|
| | NEG-J-45 | Negative | 7 | 0 | 7 | NA | NEGATIVE |
| | NEG-J-46 | Negative | 30 | 1 | 29 | NA | NEGATIVE |
| | NEG-J-47 | Negative | 33 | 3 | 30 | NA | NEGATIVE |
| | NEG-J-48 | Negative | 12 | 0 | 12 | NA | NEGATIVE |
| | NEG-J-49 | Negative | 24 | 1 | 23 | NA | NEGATIVE |
| | NEG-J-50 | Negative | 14 | 0 | 14 | NA | NEGATIVE |
| | NEG-J-51 | Negative | 22 | 0 | 22 | NA | NEGATIVE |
| | NEG-J-52 | Negative | 8 | 0 | 8 | NA | NEGATIVE |
| | NEG-J-53 | Negative | 39 | 1 | 38 | NA | NEGATIVE |
| | NEG-J-55 | Negative | 26 | 0 | 26 | NA | NEGATIVE |
| | NEG-J-56 | Negative | 16 | 1 | 15 | NA | NEGATIVE |
| | NEG-J-57 | Negative | 18 | 2 | 16 | NA | NEGATIVE |
| | NEG-J-58 | Negative | 29 | 4 | 25 | NA | NEGATIVE |
| | NEG-J-59 | Negative | 1068 | 298 | 770 | NA | NEGATIVE |
| | Neg-J-60 | Negative | 267 | 54 | 213 | NA | NEGATIVE |
| | NEG-J-61 | Negative | 1307 | 377 | 930 | NA | NEGATIVE |
| | Neg-J-62 | Negative | 1419 | 384 | 1035 | NA | NEGATIVE |
| | SC2-A-01 | SARS-CoV-2 | 2492 | 1654 | 838 | 0 | POSITIVE |
| | SC2-A-02 | SARS-CoV-2 | 5024 | 4663 | 361 | 0 | POSITIVE |
| | SC2-A-03 | SARS-CoV-2 | 2214 | 806 | 1408 | 3.83E-08 | POSITIVE |
| | SC2-A-04 | SARS-CoV-2 | 490 | 172 | 318 | 0.049608197 | NEGATIVE |
| | SC2-A-05 | SARS-CoV-2 | 931 | 826 | 105 | 0 | POSITIVE |
| | SC2-A-06 | SARS-CoV-2 | 4118 | 4045 | 73 | 0 | POSITIVE |
| | SC2-A-07 | SARS-CoV-2 | 127 | 99 | 28 | 2.61E-30 | POSITIVE |
| | SC2-A-08 | SARS-CoV-2 | 771 | 559 | 212 | 4.81E-137 | POSITIVE |
| | SC2-A-09 | SARS-CoV-2 | 3265 | 3252 | 13 | 0 | POSITIVE |
| | SC2-A-10 | SARS-CoV-2 | 313 | 144 | 169 | 9.44E-09 | POSITIVE |
| | SC2-A-11 | SARS-CoV-2 | 1367 | 494 | 873 | 4.01E-05 | POSITIVE |
| | SC2-A-12 | SARS-CoV-2 | 68 | 38 | 30 | 9.14E-06 | POSITIVE |
| | SC2-D-01 | SARS-CoV-2 | 8351 | 8342 | 9 | 0 | POSITIVE |
| | SC2-D-03 | SARS-CoV-2 | 1191 | 945 | 246 | 5.65E-285 | POSITIVE |
| | SC2-D-04 | SARS-CoV-2 | 2315 | 2159 | 156 | 0 | POSITIVE |
| | SC2-D-09 | SARS-CoV-2 | 348 | 266 | 82 | 5.04E-75 | POSITIVE |
| | SC2-D-10 | SARS-CoV-2 | 324 | 81 | 243 | NA | NEGATIVE |
| | SC2-D-12 | SARS-CoV-2 | 1163 | 455 | 708 | 2.10E-09 | POSITIVE |
| | SC2-D-13 | SARS-CoV-2 | 660 | 236 | 424 | 0.008224134 | POSITIVE |
| | SC2-D-25 | SARS-CoV-2 | 337 | 126 | 211 | 0.011217453 | POSITIVE |
| | SC2-D-28 | SARS-CoV-2 | 1510 | 771 | 739 | 9.79E-64 | POSITIVE |
| | SC2-D-29 | SARS-CoV-2 | 1347 | 1052 | 295 | 9.54E-306 | POSITIVE |
| | SC2-WT-01 | SARS-CoV-2 | 4058 | 3895 | 163 | 0 | POSITIVE |
| | SC2-WT-02 | SARS-CoV-2 | 1609 | 1149 | 460 | 4.10E-269 | POSITIVE |
| | SC2-WT-03 | SARS-CoV-2 | 1281 | 1123 | 158 | 0 | POSITIVE |
| | SC2-WT-04 | SARS-CoV-2 | 1206 | 830 | 376 | 2.02E-177 | POSITIVE |
| | SC2-WT-05 | SARS-CoV-2 | 2043 | 1740 | 303 | 0 | POSITIVE |
| | SC2-WT-06 | SARS-CoV-2 | 1332 | 1264 | 68 | 0 | POSITIVE |
| | SC2-WT-07 | SARS-CoV-2 | 2777 | 2736 | 41 | 0 | POSITIVE |
| | SC2-WT-08 | SARS-CoV-2 | 860 | 437 | 423 | 3.35E-36 | POSITIVE |
| | SC2-WT-09 | SARS-CoV-2 | 2130 | 2126 | 4 | 0 | POSITIVE |
| | SC2-WT-10 | SARS-CoV-2 | 1600 | 1411 | 189 | 0 | POSITIVE |
| | SC2-WT-11 | SARS-CoV-2 | 1671 | 1311 | 360 | 0 | POSITIVE |
| | SC2-WT-12 | SARS-CoV-2 | 2301 | 1862 | 439 | 0 | POSITIVE |
| | SC2-WT-13 | SARS-CoV-2 | 2808 | 2252 | 556 | 0 | POSITIVE |
| | SC2-WT-14 | SARS-CoV-2 | 4295 | 4286 | 9 | 0 | POSITIVE |

**Table 4. Results of variant classification (35 samples).** Abbreviations used: Neg – negative, NA – statistical test not needed as the percentage of BBXs classified as positive by the trained network (i.e. # positives/total # of BBXs) for either variant is greater than the sensitivity for said variant, then the sample is classified as said variant, SC2 – SARS-CoV-2. Samples with p-values closer than 3 orders of magnitude (i.e. inconclusive) are highlighted in yellow and incorrectly classified samples are highlighted in turquoise.

| **Network** | **Identifier** | **RT-PCR** | **BBXs breakdown** | | | **P-value** | | **Result** |
|---|---|---|---|---|---|---|---|---|
| 'Microscope 3' | | | Total # of BBXs | A+WT | D | P-A+WT<br>Null hypothesis A+WT | P-D<br>Null hypothesis D | |
| **SARS-CoV-2 Alpha+Wuhan vs SARS-CoV-2 Delta** | NEG-J-29 | Negative | 508 | 325 | 183 | 9.58385E-09 | 1.30046E-21 | Delta |
| | SC2-D-01 | Delta Variant | 8342 | 409 | 7933 | 0 | NA | Delta |
| | SC2-D-03 | Delta Variant | 31 | 30 | 1 | NA | 1.47E-09 | Alpha-Wuhan |
| | SC2-D-04 | Delta Variant | 9 | 4 | 5 | 0.03426401 | 0.930251279 | Inconclusive |
| | SC2-D-09 | Delta Variant | 266 | 133 | 133 | 4.67E-21 | 0.021108289 | Delta |
| | SC2-D-12 | Delta Variant | 455 | 223 | 232 | 1.59E-37 | 0.009600884 | Delta |
| | SC2-D-13 | Delta Variant | 236 | 128 | 108 | 1.76E-13 | 0.000488561 | Delta |
| | SC2-D-25 | Delta Variant | 337 | 109 | 228 | 4.27E-73 | NA | Delta |
| | SC2-D-28 | Delta Variant | 771 | 549 | 222 | 0.01498451 | 2.27E-56 | Alpha-Wuhan |
| | SC2-D-29 | Delta Variant | 1052 | 184 | 868 | 0 | NA | Delta |
| | SC2-S0-01 | Wuhan/Alpha Variant | 1654 | 1248 | 406 | NA | 1.38E-156 | Alpha-Wuhan |
| | SC2-S0-02 | Wuhan/Alpha Variant | 4663 | 4513 | 150 | NA | 0 | Alpha-Wuhan |
| | SC2-S0-03 | Wuhan/Alpha Variant | 806 | 672 | 134 | NA | 1.36E-118 | Alpha-Wuhan |
| | SC2-S0-05 | Wuhan/Alpha Variant | 526 | 476 | 50 | NA | 2.75E-107 | Alpha-Wuhan |
| | SC2-S0-06 | Wuhan/Alpha Variant | 4045 | 3530 | 515 | NA | 0 | Alpha-Wuhan |
| | SC2-S0-07 | Wuhan/Alpha Variant | 99 | 89 | 10 | NA | 4.28E-21 | Alpha-Wuhan |
| | SC2-S0-08 | Wuhan/Alpha Variant | 559 | 529 | 30 | NA | 2.99E-134 | Alpha-Wuhan |
| | SC2-S0-09 | Wuhan/Alpha Variant | 3252 | 3051 | 201 | NA | 0 | Alpha-Wuhan |
| | SC2-S0-10 | Wuhan/Alpha Variant | 144 | 72 | 72 | 4.26219E-12 | 0.089751493 | Delta |
| | SC2-S0-11 | Wuhan/Alpha Variant | 494 | 147 | 347 | 2.68E-119 | NA | Delta |
| | SC2-S0-12 | Wuhan/Alpha Variant | 38 | 22 | 16 | 0.01488672 | 0.063651707 | Inconclusive |
| | SC2-WT-01 | Wuhan/Alpha Variant | 3895 | 3740 | 155 | NA | 0 | Alpha-Wuhan |
| | SC2-WT-02 | Wuhan/Alpha Variant | 1149 | 868 | 281 | NA | 5.50E-110 | Alpha-Wuhan |
| | SC2-WT-03 | Wuhan/Alpha Variant | 1123 | 1025 | 98 | NA | 3.45E-234 | Alpha-Wuhan |
| | SC2-WT-04 | Wuhan/Alpha Variant | 830 | 770 | 60 | NA | 1.92E-184 | Alpha-Wuhan |
| | SC2-WT-05 | Wuhan/Alpha Variant | 1740 | 1541 | 199 | NA | 0 | Alpha-Wuhan |
| | SC2-WT-06 | Wuhan/Alpha Variant | 1264 | 1190 | 74 | NA | 2.51E-295 | Alpha-Wuhan |
| | SC2-WT-07 | Wuhan/Alpha Variant | 2736 | 2516 | 220 | NA | 0 | Alpha-Wuhan |
| | SC2-WT-08 | Wuhan/Alpha Variant | 860 | 501 | 359 | 8.32E-30 | 1.61E-19 | Delta |
| | SC2-WT-09 | Wuhan/Alpha Variant | 2126 | 574 | 1552 | 0 | NA | Delta |

| | | | | | | | | |
|---|---|---|---|---|---|---|---|---|
| | SC2-WT-10 | Wuhan/Alpha Variant | 1411 | 1317 | 94 | NA | 0 | Alpha-Wuhan |
| | SC2-WT-11 | Wuhan/Alpha Variant | 1311 | 1221 | 90 | NA | 2.49E-294 | Alpha-Wuhan |
| | SC2-WT-12 | Wuhan/Alpha Variant | 1862 | 1590 | 272 | NA | 7.56E-299 | Alpha-Wuhan |
| | SC2-WT-13 | Wuhan/Alpha Variant | 2252 | 1895 | 357 | NA | 0 | Alpha-Wuhan |
| | SC2-WT-14 | Wuhan/Alpha Variant | 4286 | 3696 | 590 | NA | 0 | Alpha-Wuhan |
